# Comparison of B-cell depletion versus natalizumab for treatment of multiple sclerosis: A semi-supervised causal analysis

**DOI:** 10.1101/2025.01.24.25321100

**Authors:** Dominic DiSanto, Weijing Tang, Wen Zhu, Michele Morris, Ratnam Srivastava, Shyam Visweswaran, Tianxi Cai, Zongqi Xia

**Author notes:** Share co-first authorship (Dominic DiSanto, Weijing Tang). Share co-senior authorship (Tianxi Cai, Zongqi Xia).

## Abstract

**Background:** B-cell depletion (BCD) therapies (*e.g.,* ocrelizumab, ofatumumab, rituximab) and natalizumab (NTZ) are highly effective disease-modifying therapies (DMTs) for multiple sclerosis (MS). However, no randomized clinical trial and only limited observational studies compared the two DMT classes.

**Objective:** We compared BCD and NTZ in managing MS patient-reported disability progression using registry-linked electronic healthcare record (EHR) data.

**Methods:** The study population of an EHR cohort of MS patients included a subset enrolled in a clinic-based MS registry that provided gold-standard outcome labels. To estimate average treatment effects, we applied a doubly-robust semi-supervised approach to analyze all (not only registry) patients and comprehensively adjusted for confounders that included not only *a priori* standard features but also knowledge graph-derived EHR features. While gold-standard disability outcomes were available in registry patients, we imputed the baseline pre-treatment and post-treatment disability status for non-registry patients. We categorized patient-reported disability progression status as “sustained worsening”, “sustained improvement”, or “no sustained change” based on 3 or more observations or imputations of Patient Determined Disease Steps (PDDS) scores within 3 years after target treatment initiation as the primary endpoint.

**Results:** In this MS cohort (n=1,738, Age=46±13 years, Non-Hispanic White=86.71%), there was no significant difference between BCD (n=1,245, 71.63%) and NTZ (n=495, 28.37%) in mitigating sustained worsening (ATE=-0.020, 95% CI [-0.149, 0.076], p=.755) or promoting sustained improvement (ATE=-0.073, 95% CI [-0.187, 0.009], p=.114) of patient-reported disability. Sensitivity analyses using a 2-year window after treatment initiation confirmed no difference in sustained worsening (ATE=-0.013, 95% CI [-0.069, 0.074], p=.819) or sustained improvement (ATE=-0.187, 95% CI [-0.264, 0.008], p=.135) between BCD and NTZ. In power analysis, the semi-supervised approach increased statistical power compared to the standard approach of using gold-standard data alone.

**Conclusion:** This real-world comparative effectiveness analysis based on a novel doubly-robust semi-supervised approach found no difference between BCD and NTZ in managing MS disability progression.

**Key Messages:**

▪ Evaluation of sustained disability accumulation requires long-term follow-up beyond the typical clinical trials, while the scarcity of patient-reported and certainly rater-assessed disability outcomes in routine clinical care hinders analysis using real-world clinical data.
▪ Using a large registry-linked electronic healthcare record cohort and a novel semi-supervised, doubly-robust method that incorporates knowledge graph-derived clinically relevant covariates from EHR, we conducted a causal inference study to compare sustained change in patient-reported disability in people with MS.
▪ The semi-supervised approach effectively leverages additional data from patients without observed outcome information and increases the statistical power of the comparative effectiveness study while retaining robustness properties and achieving more consistent treatment effect estimation in the causal analysis.
▪ There was no statistically significant difference between B-cell depletion therapy and natalizumab in sustained patient-reported disability outcomes up to 3-years after treatment initiation.

## Introduction

Multiple sclerosis (MS) is characterized by inflammatory demyelination and progressive neurodegeneration in the central nervous system. Both neuroinflammation (*i.e.,* relapses) and neurodegeneration contribute to neurological disability accumulation in people with MS (pwMS). Disease-modifying therapies (DMTs) have improved MS outcomes by reducing relapses and delaying disability worsening. Among common DMTs in the United States (US), B-cell depletion (BCD) therapies (*e.g.,* ocrelizumab, ofatumumab, rituximab) and natalizumab (NTZ) are associated with reduced disability worsening in pwMS.^1–10^ However, no randomized clinical trials (RCTs) and limited real-world studies have directly compared the effectiveness of these DMTs in managing disability.^1,3–6,8–11^

The clinician-assessed Expanded Disability Status Scale (EDSS) is the most recognized disability measure in MS. While commonly used in RCTs, EDSS is impractical for routine monitoring or research collection due to cost (*e.g.,* need for trained personnel) or patient burden. In contrast, the Patient Determined Disease Steps (PDDS) scale is a validated patient-reported disability measure that is significantly easier to administer and has been increasingly adopted in clinical practice.^12^

While registries collect outcomes unavailable in routine patient care, electronic health records (EHRs) provide longitudinal clinical data. These complementary data sources have enabled integrative analyses of data from MS registry and EHR data to model disease severity,^13^ identify comorbidities associated with MS severity,^14^ predict future relapse,^15^ evaluate temporal trends in relapse rate,^16^ and adjust for confounders when comparing DMTs for relapse reduction.^17^ However, registry participants may not represent the broader patient population, potentially limiting the generalizability of registry-based findings. Real-world studies face the challenge of missingness outcomes. Advances in statistical methods have introduced doubly-robust estimation of average treatment effects (ATEs) in semi-supervised settings, enabling causal inference when a subset of patients lack outcome labels.^18,19^ Deploying semi-supervised methods, we can leverage both registry-enrolled patients and non-registry patients without outcome data. This study employs a novel, doubly-robust semi-supervised approach to assess the comparative effectiveness of two highly effective DMT classes (BCD vs NTZ) in managing patient-reported sustained disability outcomes in pwMS.

## Methods

### Ethics Approval

The University of Pittsburgh Institutional Review Board approved the study protocols (STUDY19080007, STUDY21030127). All registry participants provided written informed consent. EHR research protocol was deemed exempt.

### Data Source

We used EHR data from the University of Pittsburgh Medical Center (UPMC). Codified data (available from January 1, 2004 to December 31, 2022) contained demographic and clinical information, including daily counts of codes indicating diagnoses (*e.g.,* International Classification of Disease [ICD]), procedures (*e.g.,* current procedural terminology [CPT]), prescriptions (*e.g.,* RxNorm), and laboratory tests (*e.g.,* Logical Observation Identifiers Names and Codes [LOINC]). We mapped all ICD codes to PheCodes,^20^ consolidated CPT procedure codes using the Clinical Classifications Software (CCS) for Services and Procedures, and grouped electronic prescription codes to the RxNorm ingredient level (see **Supplementary Materials-EHR Data Pre-processing**). Narrative data (free-text clinical narratives available from January 1, 2011 to December 31, 2022) were processed using a previously validated natural language processing pipeline, leveraging the unified medical language system, to generate concept unique identifiers (CUIs).^21,22^

We used EHR-linked registry data of a subset of patients enrolled in a clinic-based cohort, Prospective Investigation of Multiple Sclerosis in the Three Rivers Region (PROMOTE, Pittsburgh, PA), between 2017 and 2023. Registry data included longitudinal outcomes (*i.e.,* PDDS) as well as DMT records, which were integrated with prescriptions from EHR to assign treatment exposure arms.

### Cohort Derivation

The cohort follow-up window (2014-2022) balances sufficient sample size while mitigating temporal shifts in clinical practice. We used the Knowledge-Driven Online Multimodal Automated Phenotyping (KOMAP) algorithm^23^ to identify patients with an MS diagnosis. Among 17,827 potentially eligible patients, 3,039 pwMS received their first target treatment (BCD or NTZ) between January 1, 2014, and December 31, 2022. We excluded 993 patients who had received other highly effective DMTs (*e.g.,* alemtuzumab, cladribine, mitoxantrone) or target treatment prior to January 1, 2014, as well as 76 patients who ever received chemotherapies (*e.g.,* cyclophosphamide, methotrexate). We further excluded 217 patients who switched DMTs after target treatment initiation. Finally, we excluded 15 patients with missing demographic or clinical features. This final cohort included 1,738 MS patients, including 667 registry-enrolled participants.

### Treatment Assignment

We compared two highly effective DMT mechanistic classes: BCD (*i.e.,* ocrelizumab, ofatumumab, rituximab) and NTZ. Although not officially approved for MS, rituximab is widely used in clinical practice. Ublituximab was excluded due to its approval after the data extraction date. We assigned a patient’s treatment exposure arm if first receiving target treatment between January 1, 2014 and December 31, 2022. The date of first receiving BCD or NTZ was the target treatment initiation date.

### Disability Outcomes

We assessed sustained changes in disability based on PDDS^11,24,25^, which is a validated scale of patient-reported disability in MS. PDDS scores range from 0 to 8, indicating increasing disability. Due to sparse observations of scores of 8 (<1%), we combined scores of 7 and 8 into a single category.

We categorized patients into four distinct groups: “sustained worsening”, “sustained improvement”, “no sustained change”, and “insufficient information”. Adjudication of *sustained* changes required at least three PDDS scores, separated by at least 6 months between consecutive scores, within 3 years after treatment initiation. Sustained worsening or improvement required two sequential PDDS scores that were either greater or lesser, respectively, compared to the first observed PDDS score after treatment initiation. We considered the first occurrence of sustained change as the outcome (*i.e.,* worsening or improvement, whichever occurred earlier). We referred to patients with “insufficient information” as having “***unlabeled outcome***” or “***no labeled outcome****”*, and those with the other three categories as having “***labeled***” outcomes. Labeled outcome data were available only in a subset of registry-enrolled participants.

We analyzed sustained worsening and sustained improvement separately. When evaluating sustained worsening, we merged the other categories of labeled patients as “no sustained worsening”: *i.e.,* patients with “sustained improvement” and “no sustained change”. When evaluating sustained improvement, we likewise merged the other two labeled categories as “no sustained improvement”.

### Confounders

To control potential confounding, we adjusted for baseline features based on standard clinical trial practice, healthcare utilization, and knowledge-graph derived features from EHR. We obtained the covariates primarily from the EHR and supplemented with registry data. Standard clinical trial features included demographics, disease duration (*i.e.,* from first MS PheCode to target treatment initiation), follow-up duration (*i.e.,* from first clinical encounter to treatment initiation), prior DMT use duration (*i.e.,* from earliest record of prior DMT use to treatment initiation; or 0 for those without prior DMT).

As disability status at treatment initiation was infrequently available, we calculated a baseline risk covariate for all patients using a PDDS imputation model based on pre-target treatment clinical history. Imputation models were developed using a separate dataset of PDDS scores unused in this causal analysis. See **Supplementary Materials**-**PDDS Imputation Model, Supplementary Tables 1a-b, 2; Supplementary** Figure 1) for details on imputation models and performance.

Healthcare utilization features indicate not only overall health status but also potentially missing information in the fragmented US healthcare landscape. To capture *total* baseline healthcare utilization, we included a covariate of the counts of codified EHR features and narrative CUI’s that were observed during distinct clinical encounters, occurring prior to target treatment initiation. We included a covariate of the counts of distinct clinical encounters with an MS PheCode (based on distinct code-date pairings).

To pre-select EHR features, we used an online narrative and codified feature search engine (ONCE), powered by pre-trained multi-source knowledge graph representation learning of EHR concepts.^23^ See detailed methods in **Supplementary Materials-Knowledge Graph-Derived Feature Selection** and the included EHR features in **Supplementary Tables 3a-d**. To address feature sparsity, we excluded features with <10% frequency in the cohort. We used normalized aggregated counts of diagnoses (PheCodes), procedures (CCS codes), prescriptions (RxNorm codes), laboratory tests (LOINC codes), and clinical narrative features (CUIs) before treatment initiation, dividing the total occurrence of each feature by patient-level total healthcare utilization.

### Statistical Analysis

We employed a doubly-robust, semi-supervised estimation method for causal analysis.^26,27^ To assess causal effect using observational data, it is crucial to adjust for confounders associated with treatment assignment and sustained change outcome. Doubly-robust methods yield consistent causal estimates despite misspecification in either the treatment or outcome model. In contrast to the standard methods that rely solely on labeled data, this semi-supervised method incorporates both labeled and imputed outcome labels. The study design is presented in **Figure 1**.

**Figure 1.**
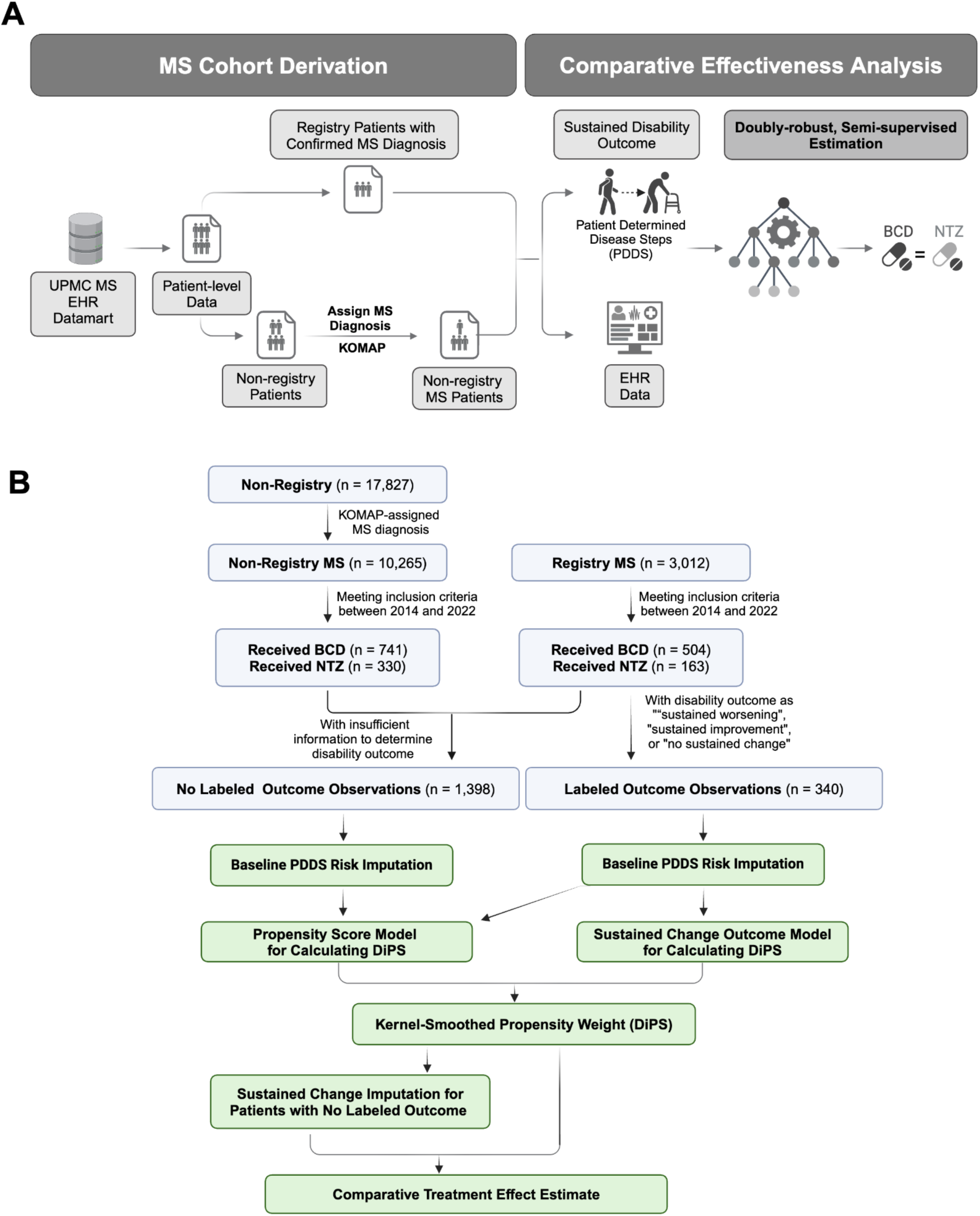
Study design. (A) Schematic overview. (B) Flowchart of cohort derivation steps (boxes with blue outline) and the modeling steps in the causal analysis (green boxes). The “Non-Registry - MS” panel represents patients with a KOMAP algorithm-phenotyped MS diagnosis who were not enrolled in the clinic-based MS registry. The “Registry - MS” panel represents patients with neurologist-confirmed MS diagnosis who were enrolled in the clinic-based MS registry (with linked EHR data). Sample sizes for the “Received” DMT categories correspond to patients with a first record of BCD or NTZ between January 1, 2014 and December 31, 2022. See Methods for details on the cohort derivation. “Labeled outcome observations” correspond to patients with “sustained worsening”, “sustained improvement”, or “no sustained change” in PDDS score, while “no labeled outcome observations” correspond to patients with “insufficient information” for assessing sustained changes. **Abbreviations:** BCD, B-Cell Depletion therapy. DiPS, Double-Index Propensity Score. DMT, Disease Modifying Therapy. EHR, Electronic Health Record. KOMAP, Knowledge-Driven Online Multimodal Automated Phenotyping. MS, Multiple Sclerosis. NTZ, Natalizumab. PDDS, Patient Determined Disease Steps. UPMC, University of Pittsburgh medical Center.

The semi-supervised estimation consists of three steps. First, we fit initial propensity and outcome regression adaptive Least Absolute Shrinkage and Selection Operator (LASSO) models to calculate a modified version of a treatment propensity weight, specifically a kernel-smoothed propensity score, *i.e.* the **double-index propensity score (DiPS)**. Second, we developed an outcome imputation model that predicts sustained-change outcomes. Finally, we estimate the treatment effect using a **doubly-robust inverse propensity weighting** (IPW) estimator, using the DiPS as balancing weights and incorporating imputed outcome labels.^26,27^ This doubly-robust IPW estimator ensures consistency under correct specification of either an initial propensity score model or initial outcome model. Additionally, it is robust to misspecification of the outcome imputation model and generates consistent estimates.

The outcome imputation model for sustained change in the second step requires additional covariates. Specifically, this model retains the same expert-defined codified and narrative features from the initial outcome model, extended to include aggregated data up to one year after treatment initiation. Additionally, we added three new covariates: a binary variable of target treatment group membership, an inverse DiPS weight for the assigned treatment, and a third capturing PDDS risk at one year after treatment initiation. For patients with observed PDDS scores within the first year after treatment initiation, we used the average of their scores, while we estimated their scores using an independent PDDS imputation model based on codified and narrative data from the first year for those without observed PDDS scores. Finally, we trained the outcome imputation model using LASSO logistic regression on labeled data and applied it to impute the sustained change outcome when a labeled outcome was unavailable.

The final comparative treatment effect estimation between BCD and NTZ was based on DiPS-weighted outcomes on all patients, using the observed outcomes for labeled data and imputed outcomes in the absence of labeled outcome. We employed a perturbation resampling procedure to construct confidence intervals for estimates and calculated two-sided p-values through confidence interval inversion.^27,28^ We considered p-values below 0.05 as statistically significant. Further, we conducted a power evaluation based on a normal approximation, using the standard error estimated from perturbation analysis. See details in **Supplementary Materials-Detailed Causal Analysis**.

### Sensitivity Analyses

Data pre-processing prior to the main analysis included selecting a time-window for pre-treatment feature calculation, prevalence cut-off for EHR features trimming, window selection for post-treatment PDDS scores to ascertain the sustainment outcome, knowledge graph screening of EHR features, and exclusion of patients who switched DMTs during the study period. To validate the main findings, we performed sensitivity analyses, altering each of these analytic decisions. We repeated the analyses using 6- and 12-month pre-treatment periods, applying a 5% prevalence threshold for EHR features, categorizing outcome status with a 3-month window between PDDS scores, using all observed EHR features rather than knowledge graph-derived subset, and including individuals who switched DMT’s during the study period. The inclusion of DMT switchers mirrors an intention-to-treat analysis. We repeated all analyses using 2- and 3-year endpoints.

### Data Availability

We will publicly disseminate anonymous summary-level registry data and EHR data. The rationale for not sharing patient-level data is that patient-level clinical data (either de-identified information or limited protected health information containing dates of clinical events or even if anonymous due to concern for re-identification) are universally subject to the rules and regulation of each healthcare system, which may only be affiliated with but are not the same as the primary academic institutions of the study investigators. Sharing of de-identified EHR data with qualified external researchers by each of the study performance site may be permissible only after the approval of the respective Institutional Review Boards (IRBs), regulatory oversight agents of the healthcare systems (that own the clinical data) as well as the appropriate Data Usage Agreements (DUA) between institutions.

Code for analysis and figures is available at: <https://github.com/xialab2016/BCD_NTZ_SemiSupervisedCausal>.

## Results

### Patient Characteristics

**Table 1** displays characteristics comparing registry and non-registry patients. Registry-enrolled patients demonstrate higher mean total baseline healthcare utilization. In the overall study cohort, 1,245 (71.63%) patients received BCD, while 1,398 (80.44%) did not have labeled sustained-change outcomes. **Figure 2** displays the relative average or proportion of characteristics by treatment class and by sustained change outcome category as compared to the overall mean. When compared to patients in the NTZ group, patients in the BCD group had longer disease duration (5.62 vs 4.33 years), follow-up duration (9.82 vs 8.05 years), higher mean total baseline healthcare utilization (1,561 vs 900 distinct codes and CUIs), higher baseline PDDS risk (2.13 vs 1.88), and fewer women (68.35% vs 80.93%). Patients with sustained disability improvement had lower baseline and year-1 PDDS risk and were more often women. Additional summaries by outcome category and treatment group are available in **Supplementary Tables 4a-c**. We compared baseline covariates after balancing by our DiPS weights, which reached appropriate balance by target treatment class (**Supplementary Tables 5a-b**).

**Figure 2.**
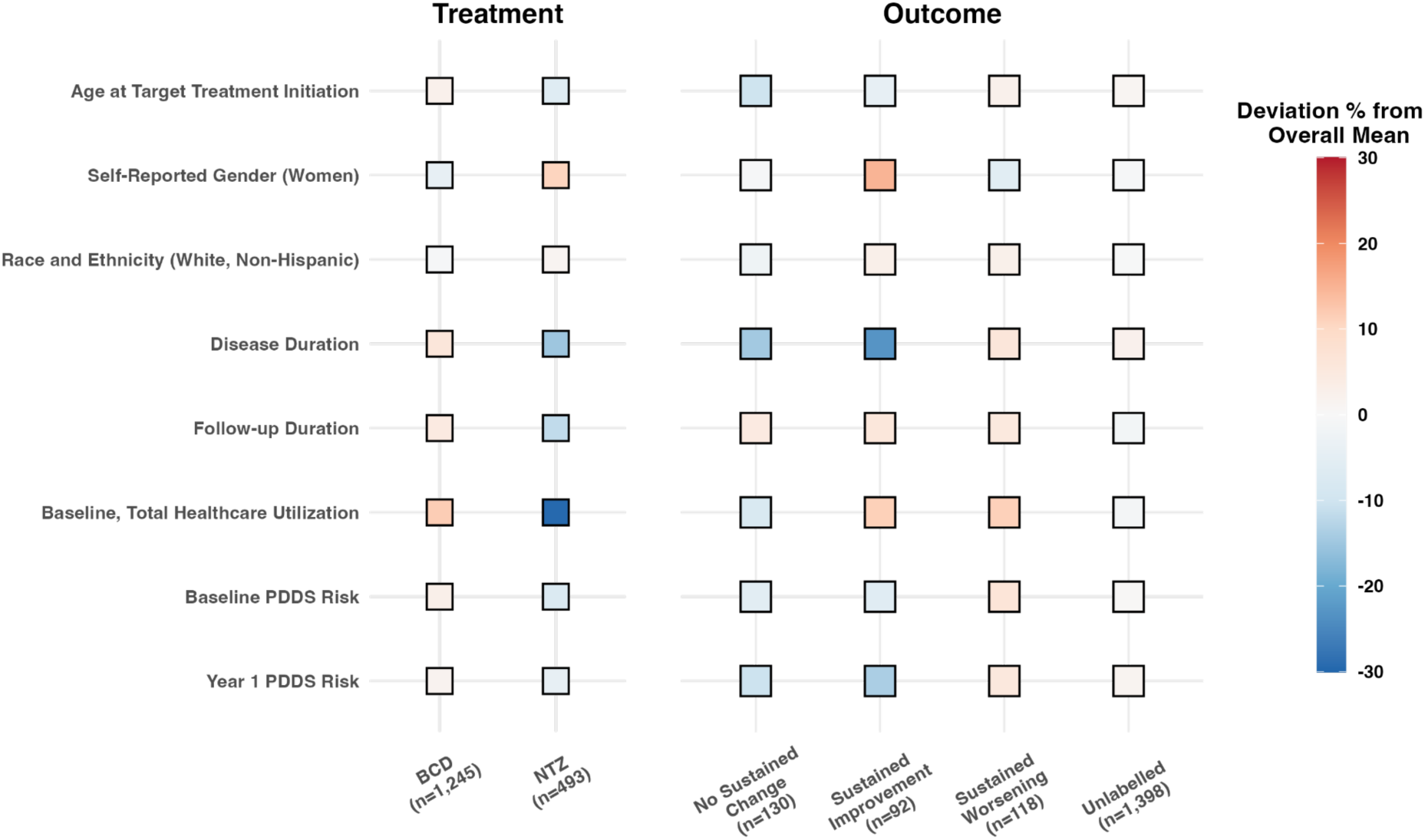
**Cohort characteristics by treatment class and outcome category.** Each square reports the difference between the column-wise subgroup mean and the overall cohort mean for the variable listed in each row. Red squares indicate values for a sub-group being greater than the overall mean, while blue squares indicate values being less than the overall mean. The left panel summarizes differences across treatment assignments. The right panel summarizes differences across the four sustained change categories. For categorical variables (Women, White, Non-Hispanic race/ethnicity), the mean corresponds to the percentage of the category. **Abbreviations**: BCD, B-Cell Depleting drugs. NTZ, Natalizumab. PDDS, Patient Determined Disease Steps.

**Table 1.**
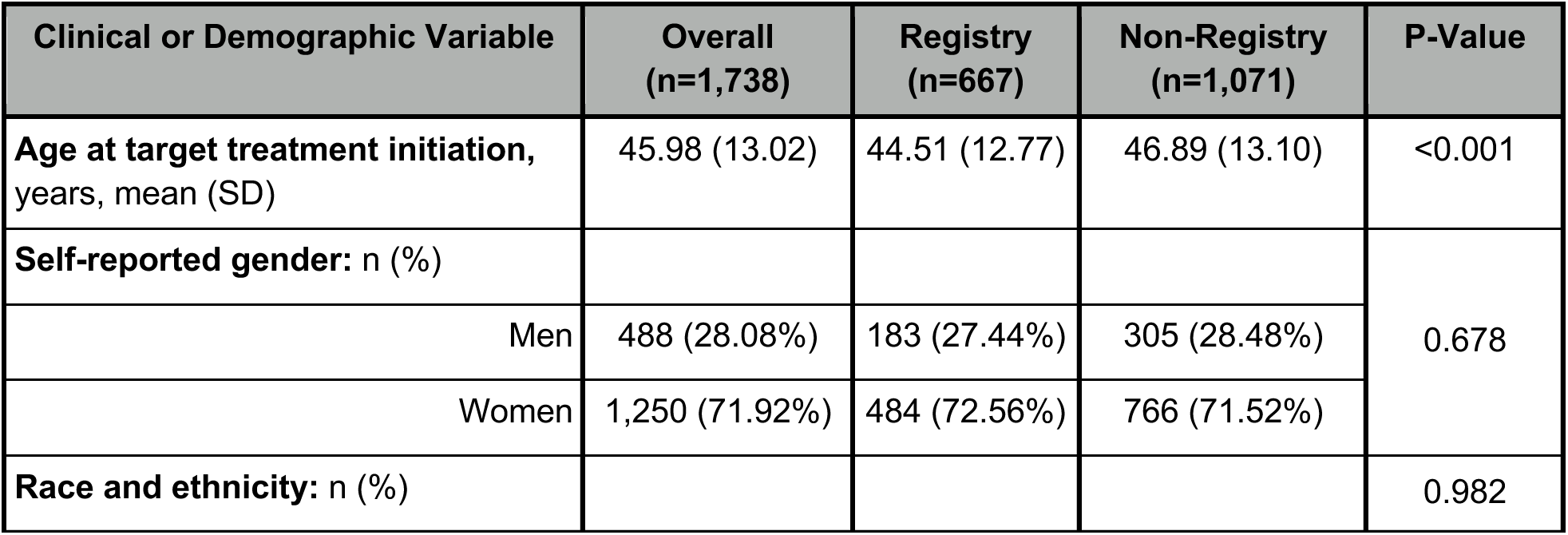
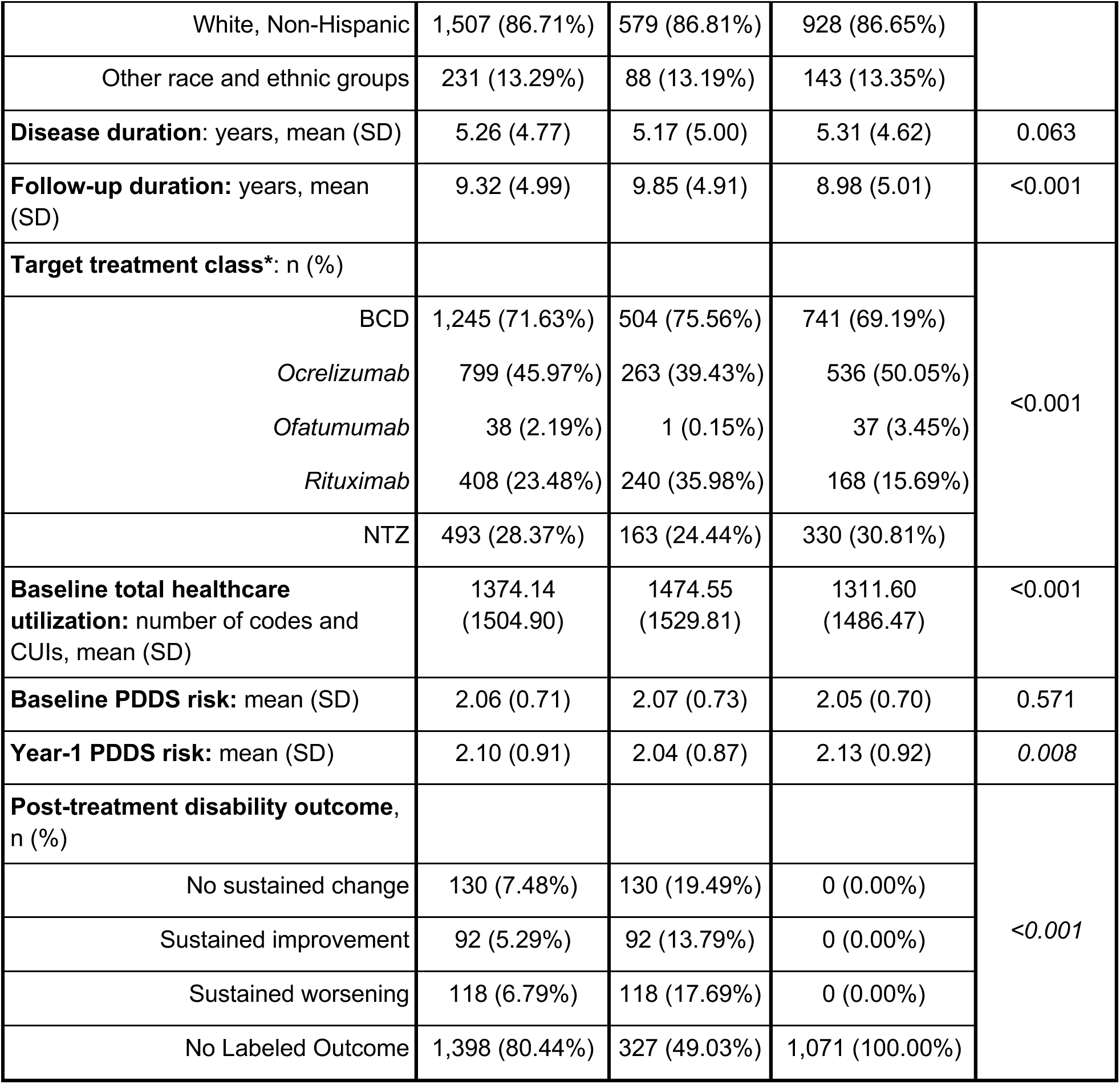
Cohort characteristics. **Note:** We presented mean (standard deviation) for continuous variables and count (percentage) for categorical variables. Baseline total healthcare utilization was the count of all codified EHR features and narrative CUI features observed during distinct clinical encounters occurring prior to target treatment initiation. Baseline PDDS risk was derived as the predicted mean PDDS score at the time of target treatment initiation, using independently fitted PDDS imputation models. Year-1 PDDS risk was calculated as the average of observed PDDS scores within 1 year post-treatment initiation for patients with any available PDDS data and otherwise imputed by the same, independent PDDS imputation models using additional covariate information through 1 year post-treatment initiation. P-values were reported from chi-square tests for categorical variables (race-ethnicity, gender, target treatment category, and disability outcome) and Mann-Whitney U-tests for the remaining, continuous variables, comparing Registry and Non-Registry observations. (*) We reported the sample size (and the percentage within the BCD mechanistic class) of each specific BCD DMT agent. **Abbreviations**: BCD, B-cell depletion. CUI, Concept Unique identifier. DMT, Disease Modifying Therapy. NTZ, Natalizumab. PDDS, Patient Determined Disease Steps.

### Comparative Effectiveness Analysis

**Table 2** contains the causal analysis results. Higher PDDS scores indicate greater patient-reported disability. With NTZ as the reference treatment class, a positive ATE for the sustained worsening outcome would indicate BCD as *less* effective than NTZ in reducing patient-reported disability worsening. A positive ATE for sustained improvement outcome would indicate BCD as *more* effective than NTZ. We observed no statistically significant difference between BCD and NTZ for either sustainment outcome.

**Table 2.** Comparative treatment effect with NTZ as the reference treatment class. **Note:** Visualization of the results is presented in **Supplementary** Figure 2. **Abbreviations**: ATE, Average Treatment Effect; CI, Confidence Interval. Std. Err, Standard Error.

| Outcome | ATE | Std. Err | 95% CI | P-Value |
| --- | --- | --- | --- | --- |
| Sustained Worsening | -0.020 | 0.058 | (-0.149, 0.076) | 0.755 |
| Sustained Improvement | -0.073 | 0.052 | (-0.187, 0.009) | 0.114 |

Figure 3 displays the selected features for models of treatment assignment and sustained-change disability outcomes. The same propensity score model was used in analyses of both outcomes. In the treatment model, self-reported male gender, higher baseline total healthcare utilization, mental disorders (PheCode: 306) and older age at target treatment initiation were most associated with a higher probability of receiving BCD than NTZ. Features associated with increased risk of sustained disability worsening included dysuria (PheCode 599.3), muscle spasms (PheCode 350.1), MS distress (CUI C0231303), and distinct MS-related encounters (MS PheCode Frequency). Few features were selected in models of sustained disability improvement, with the largest coefficients observed for NTZ treatment group and metabolic diagnosis procedures (CUI C4263342). A higher number of MS-related clinical encounters (MS PheCode Frequency) and male gender were positively associated with sustained worsening and negatively with sustained improvement.

**Figure 3.**
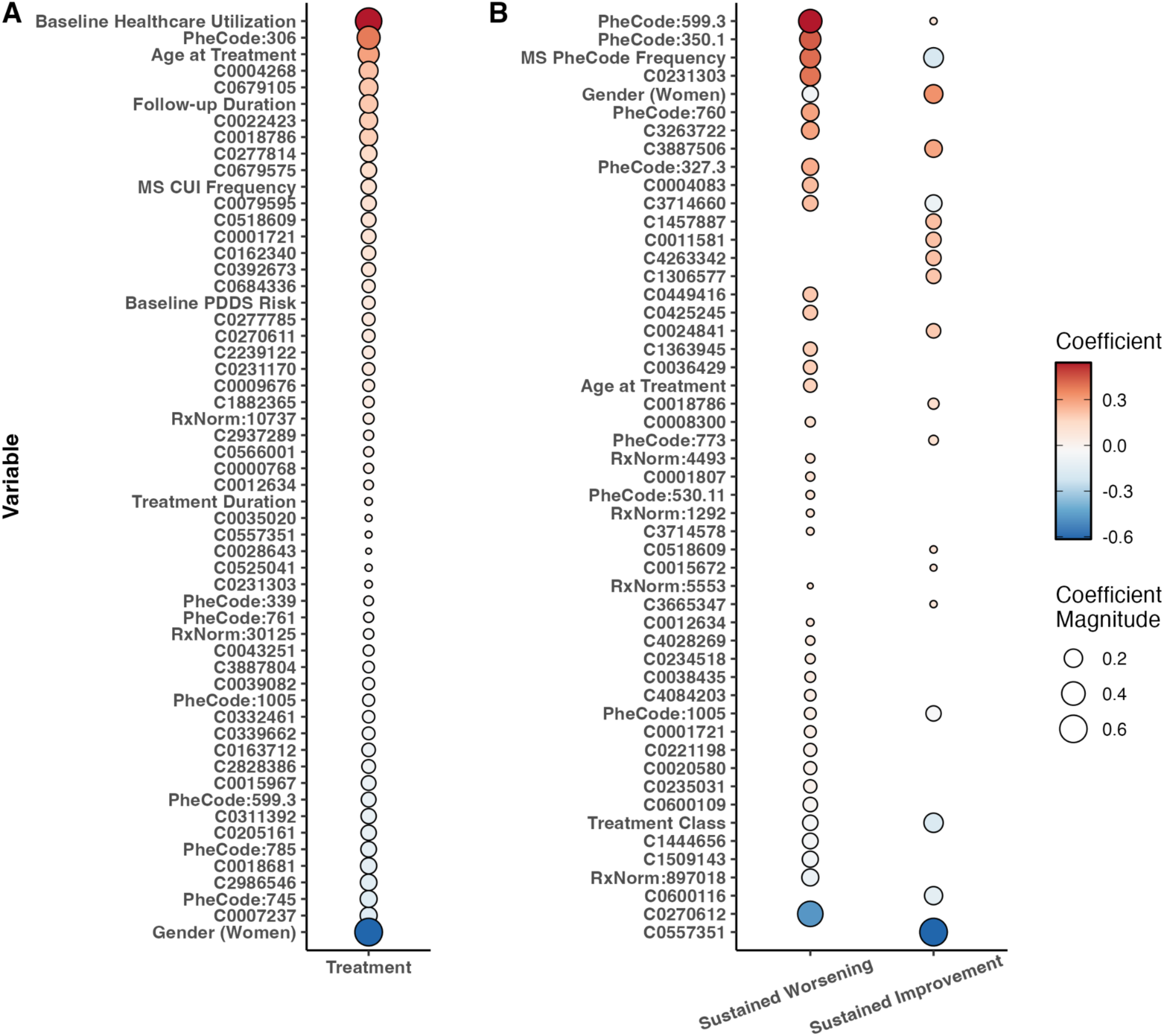
Coefficients in the treatment and outcome models for DiPS Calculation. Coefficients in the initial propensity score model (**A**) and two initial outcome models (**B**) were fitted to inform the kernel-smoothed propensity score estimator. The same treatment model was used for the final causal analysis of each outcome. These models collectively informed the final inverse probability weight that was used to weight/balance for the final ATE calculation, but no balancing/weighting was used to calculate the coefficients shown here. Only variables with non-zero coefficients were shown in each panel. Dot size represents the magnitude of the coefficient, while color corresponds to both effect size and direction. In Panel A, covariates with larger (positive) coefficients are associated with greater probability of receiving BCD than NTZ while the inverse is for smaller coefficients. Detailed descriptions of CUI, PheCode, and RxNorm codes were included in **Supplementary Tables 3a-d**. All covariates were standardized (to mean 0, unit variance) prior to model fitting. **Abbreviations:** ATE, Average Treatment Effect. BCD, B-Cell Depletion. CUI, Concept Unique Identifier. MS, Multiple Sclerosis. NTZ, Natalizumab.

### Sensitivity Analyses and Power Evaluation

Sensitivity analyses corroborated our primary results (**Supplementary** Figures 3a-b, **Supplementary Tables 6a-b**), as did analyses using a 2-year evaluation window (**Supplementary Table 7, Supplementary** Figure 2). To quantify the advantage of the semi-supervised method leveraging patients with no labeled outcome, we compared the main semi-supervised analysis to an observed-case analysis (*i.e.,* using labeled data only) (**Supplementary Tables 8a-b**). Fixing power at 80%, we reported the minimum detectable effect size for the risk difference in sustained improvement and sustained worsening between treatment groups. The semi-supervised estimate showed improvements over estimate using only observations with available outcome labels.

## Discussion

To fill knowledge gaps due to absent RCT and limited real-world evidence^29^, we applied a doubly-robust semi-supervised estimation approach to registry-linked EHR data to compare the effectiveness of two commonly prescribed DMT classes (BCD vs NTZ) in managing disability progression in pwMS. We found no statistically significant differences between BCD and NTZ in either sustained patient-reported disability outcome (worsening or improvement) up to 3-years after treatment initiation.

Prior observational studies compared the effectiveness of BCD versus NTZ in managing EDSS-based disability progression in pwMS. A retrospective study comparing ocrelizumab (n=124) and NTZ (n=157) in treatment-naive pwMS found no significant differences in time to EDSS worsening within three years after treatment initiation.^30^ Another retrospective study found no significant difference between ocrelizumab (n=310) and NTZ (n=310) in rates of confirmed EDSS progression, with median follow-up of 1.5 years.^31^ In pwMS who switched from fingolimod, a study found no difference in the rate of EDSS worsening between patients receiving ocrelizumab/rituximab (n=337) and those receiving NTZ (n=403) during a two-year follow-up.^32^ Our study reporting no significant difference between BCD and NTZ in either slowing disability worsening or promoting disability improvement complements the prior studies by evaluating BCD as a mechanistic class, assessing *sustained* disability change in *patient-reported* disability based on PDDS.

Our study has several strengths. First, our study uses a doubly-robust, semi-supervised estimation approach. Previous comparative effectiveness studies of MS DMTs that used an inverse probability of treatment weight required correct specification of the treatment model for consistent estimation of the treatment effect. In contrast, doubly-robust methods require correct specification of either the propensity score model or the outcome model, effectively increasing the likelihood of consistent treatment effect estimation. Second, the novel semi-supervised approach in this study leverages patients with and without observed outcome data from registry-linked EHR data by applying outcome imputation models to gain meaningful statistical power in the causal analysis. Third, the EHR data encompassing high-dimensional codified and narrative features provided a more comprehensive source for confounder adjustment than relying on standard covariates. Fourth, pre-trained knowledge graphs of clinical concepts coupled with regularized modelling informed the inclusion of clinically relevant features in the causal modeling while reducing feature dimensionality. Finally, the primary study outcome of sustained disability changes more reliably measures disease progression. Causal modeling with this sustainment outcome is less sensitive to possible fluctuations in this patient-reported outcome, increasing the robustness and potential generalizability of this work.

Our study also has limitations. First, while analyzing a sustained disability change outcome captures useful temporal information, this approach may overlook important temporal patterns and short-term fluctuations in disability progression within a 3-year follow-up. To partially address this concern, we conducted sensitivity analyses using a 2-year evaluation window and found consistent results. Second, while the study cohort was based in a large healthcare system and the semi-supervised estimation approach significantly augmented the statistical power, the current study (with comparable or larger sample size than prior studies comparing BCD and NTZ) might remain underpowered. Future studies involving even larger sample size, other independent cohorts, and more pragmatic rater-assessed disability outcomes than EDSS (*e.g.,* timed 25-foot walk) will be crucial to validate and further improve the generalizability of the study findings.

## Conclusion

In summary, this study contributes to the real-world evidence of the comparative effectiveness of two commonly prescribed highly effective DMT classes (BCD vs NTZ) in managing patient-reported sustained disability progression in pwMS. The doubly-robust, semi-supervised estimation approach that incorporates knowledge graph-derived clinically relevant covariates from EHR has the advantage of achieving more consistent treatment effect estimation, gaining meaningful statistical power, and adjusting relevant confounders more comprehensively than prior studies. The study findings fill the evidence gaps as there has been no RCT comparing B-cell depletion versus natalizumab and importantly create a roadmap for more robust real-world evidence generation in the future given the large number of MS DMTs for which there are no head-to-head RCT comparisons.

## References

1. Polman CH, O’Connor PW, Havrdova E, et al. A randomized, placebo-controlled trial of natalizumab for relapsing multiple sclerosis. N Engl J Med. 2006;354(9):899–910. doi:10.1056/NEJMoa044397

2. Hauser SL, Bar-Or A, Weber MS, et al. Association of Higher Ocrelizumab Exposure With Reduced Disability Progression in Multiple Sclerosis. Neurol Neuroimmunol Neuroinflammation. 2023;10(2):e200094. doi:10.1212/NXI.0000000000200094

3. Achtnichts L, Zecca C, Findling O, et al. Correlation of disability with quality of life in patients with multiple sclerosis treated with natalizumab: primary results and post hoc analysis of the TYSabri ImPROvement study (PROTYS). BMJ Neurol Open. 2023;5(1):e000304. doi:10.1136/bmjno-2022-000304

4. Rudick RA, Stuart WH, Calabresi PA, et al. Natalizumab plus interferon beta-1a for relapsing multiple sclerosis. N Engl J Med. 2006;354(9):911–923. doi:10.1056/NEJMoa044396

5. Montalban X, Hauser SL, Kappos L, et al. Ocrelizumab versus Placebo in Primary Progressive Multiple Sclerosis. N Engl J Med. 2017;376(3):209–220. doi:10.1056/NEJMoa1606468

6. Svenningsson A, Frisell T, Burman J, et al. Safety and efficacy of rituximab versus dimethyl fumarate in patients with relapsing-remitting multiple sclerosis or clinically isolated syndrome in Sweden: a rater-blinded, phase 3, randomised controlled trial. Lancet Neurol. 2022;21(8):693–703. doi:10.1016/S1474-4422(22)00209-5

7. Bar-Or A, Grove RA, Austin DJ, et al. Subcutaneous ofatumumab in patients with relapsing-remitting multiple sclerosis: The MIRROR study. Neurology. 2018;90(20):e1805–e1814. doi:10.1212/WNL.0000000000005516

8. Hauser SL, Bar-Or A, Cohen JA, et al. Ofatumumab versus Teriflunomide in Multiple Sclerosis. N Engl J Med. 2020;383(6):546–557. doi:10.1056/NEJMoa1917246

9. Hauser SL, Waubant E, Arnold DL, et al. B-cell depletion with rituximab in relapsing-remitting multiple sclerosis. N Engl J Med. 2008;358(7):676–688. doi:10.1056/NEJMoa0706383

10. Hauser SL, Bar-Or A, Comi G, et al. Ocrelizumab versus Interferon Beta-1a in Relapsing Multiple Sclerosis. N Engl J Med. 2017;376(3):221–234. doi:10.1056/NEJMoa1601277

11. Rudick RA, Lee JC, Cutter GR, et al. Disability Progression in a Clinical Trial of Relapsing-Remitting Multiple Sclerosis: Eight-Year Follow-up. Arch Neurol. 2010;67(11):1329–1335. doi:10.1001/archneurol.2010.150

12. Learmonth YC, Motl RW, Sandroff BM, Pula JH, Cadavid D. Validation of patient determined disease steps (PDDS) scale scores in persons with multiple sclerosis. BMC Neurol. 2013;13(1):37. doi:10.1186/1471-2377-13-37

13. Xia Z, Secor E, Chibnik LB, et al. Modeling disease severity in multiple sclerosis using electronic health records. PloS One. 2013;8(11):e78927. doi:10.1371/journal.pone.0078927

14. Zhang T, Goodman M, Zhu F, et al. Phenome-wide examination of comorbidity burden and multiple sclerosis disease severity. Neurol Neuroimmunol Neuroinflammation. 2020;7(6):e864. doi:10.1212/NXI.0000000000000864

15. Ahuja Y, Kim N, Liang L, et al. Leveraging electronic health records data to predict multiple sclerosis disease activity. Ann Clin Transl Neurol. 2021;8(4):800–810. doi:10.1002/acn3.51324

16. Liang L, Kim N, Hou J, et al. Temporal Trends of Multiple Sclerosis Disease Activity: Electronic Health Records Indicators. Mult Scler Relat Disord. 2022;57:103333. doi:10.1016/j.msard.2021.103333

17. Hou J, Kim N, Cai T, et al. Comparison of Dimethyl Fumarate vs Fingolimod and Rituximab vs Natalizumab for Treatment of Multiple Sclerosis. JAMA Netw Open. 2021;4(11):e2134627. doi:10.1001/jamanetworkopen.2021.34627

18. Robins JM, Rotnitzky A. Semiparametric Efficiency in Multivariate Regression Models with Missing Data. J Am Stat Assoc. 1995;90(429):122–129. doi:10.2307/2291135

19. Lunceford JK, Davidian M. Stratification and weighting via the propensity score in estimation of causal treatment effects: a comparative study. Stat Med. 2004;23(19):2937–2960. doi:10.1002/sim.1903

20. Zhang Y, Cai T, Yu S, et al. High-throughput phenotyping with electronic medical record data using a common semi-supervised approach (PheCAP). Nat Protoc. 2019;14(12):3426–3444. doi:10.1038/s41596-019-0227-6

21. Yu S, Cai T, Cai T. NILE: Fast Natural Language Processing for Electronic Health Records. Published online July 16, 2019. doi:10.48550/arXiv.1311.6063

22. Bodenreider O. The Unified Medical Language System (UMLS): integrating biomedical terminology. Nucleic Acids Res. 2004;32(Database issue):D267–D270. doi:10.1093/nar/gkh061

23. Xiong X, Sweet SM, Liu M, et al. Knowledge-Driven Online Multimodal Automated Phenotyping System. MedRxiv Prepr Serv Health Sci. Published online October 2, 2023:2023.09.29.23296239. doi:10.1101/2023.09.29.23296239

24. Salter A, Lancia S, Cutter G, et al. Characterizing Long-term Disability Progression and Employment in NARCOMS Registry Participants with Multiple Sclerosis Taking Dimethyl Fumarate. Int J MS Care. 2021;23(6):239–244. doi:10.7224/1537-2073.2020-109

25. Salter A, Lancia S, Cutter G, et al. A propensity-matched comparison of long-term disability worsening in patients with multiple sclerosis treated with dimethyl fumarate or fingolimod. Ther Adv Neurol Disord. 2021;14:17562864211021177. doi:10.1177/17562864211021177

26. Cheng D, Chakrabortty A, Ananthakrishnan AN, Cai T. Estimating average treatment effects with a double-index propensity score. Biometrics. 2020;76(3):767–777. doi:10.1111/biom.13195

27. Cheng D, Ananthakrishnan AN, Cai T. Robust and efficient semi-supervised estimation of average treatment effects with application to electronic health records data. Biometrics. 2021;77(2):413–423. doi:10.1111/biom.13298

28. Liu RY, Singh K. Notions of Limiting P Values Based on Data Depth and Bootstrap. J Am Stat Assoc. 1997;92(437):266–277. doi:10.1080/01621459.1997.10473624

29. Dahabreh IJ, Bibbins-Domingo K. Causal Inference About the Effects of Interventions From Observational Studies in Medical Journals. JAMA. 2024;331(21):1845–1853. doi:10.1001/jama.2024.7741

30. Signoriello E, Signori A, Lus G, et al. NEDA-3 achievement in early highly active relapsing remitting multiple sclerosis patients treated with Ocrelizumab or Natalizumab. Mult Scler Relat Disord. 2024;87:105594. doi:10.1016/j.msard.2024.105594

31. Boz C, Ozakbas S, Terzi M, et al. The comparative effectiveness of fingolimod, natalizumab, and ocrelizumab in relapsing-remitting multiple sclerosis. Neurol Sci Off J Ital Neurol Soc Ital Soc Clin Neurophysiol. 2023;44(6):2121–2129. doi:10.1007/s10072-023-06608-z

32. Rollot F, Couturier J, Casey R, et al. Comparative Effectiveness of Natalizumab Versus Anti-CD20 in Highly Active Relapsing-Remitting Multiple Sclerosis After Fingolimod Withdrawal. Neurother J Am Soc Exp Neurother. 2022;19(2):476–490. doi:10.1007/s13311-022-01202-1

33. Yuan Z, Zhao Z, Sun H, Li J, Wang F, Yu S. CODER: Knowledge-infused cross-lingual medical term embedding for term normalization. J Biomed Inform. 2022;126:103983. doi:10.1016/j.jbi.2021.103983

34. Crump RK, Hotz VJ, Imbens GW, Mitnik OA. Dealing with Limited Overlap in Estimation of Average Treatment Effects. Biometrika. 2009;96(1):187–199.

35. 35. Althouse AD. Post Hoc Power: Not Empowering, Just Misleading. J Surg Res. 2021;259:A3–A6. doi:10.1016/j.jss.2019.10.049

