## Supplemental Material for "Comparison of B-cell depletion versus natalizumab for treatment of multiple sclerosis: A semi-supervised causal analysis"

17

18

**Electronic Health Record (EHR) Data Pre-processing**

To systematically organize diagnoses, we mapped all ICD-9 and ICD-10 codes to PheCodes, which are manually curated to capture clinically relevant concepts for research. See further description of the PheCode pre-processing in the next paragraph. We consolidated CPT procedure codes using the Clinical Classifications Software (CCS) for Services and Procedures, a tool developed by the Agency for Healthcare Research and Quality that categorizes medical procedures into clinically meaningful groups. We grouped electronic prescription codes to the RxNorm ingredient level.
Similar to the hierarchical ICD coding taxonomy but with better curation for research, PheCodes also use sub-categories. PheCodes may include none, one, or two digits after the decimal point. As part of the pre-processing of PheCodes, we applied the following roll-up procedure to aggregate highly similar PheCodes, remove less informative codes, and reduce the overall feature dimensionality. All PheCodes with one digit after the decimal point were retained for downstream analysis. All PheCodes with two digits after the decimal point were aggregated to the one-digit level (e.g., codes 281.11, 281.12, and 281.13 would all be considered as 281.1). If a PheCode with no digit after the decimal code co-occurs with any of its one-digit "child" codes, the parent non-digit PheCode was not considered (e.g., code 345 would be excluded, if codes 345.1 and/or 345.3 were included).

**PDDS Imputation Model**

To appropriately control for all relevant confounders, the semi-supervised doubly robust IPW treatment effect estimator allows for the incorporation of non-linear effects as a covariate of pre- and post-treatment information. Among the confounders (described in the Main Methods-Confounders), we included MS disability risk as covariates in the treatment, outcome, and outcome imputation models. To estimate this latent risk, we fit imputation models for Patient Determined Disease Steps (PDDS) score, and used the imputations (in the form of predicted PDDS risk) as covariates in our causal analyses for both labeled and no labeled outcome patients. For further details, see the Supplementary Methods-Detailed Causal Analysis section.

Data for PDDS imputation model fitting included PDDS scores from patients who were operationally defined as “labeled” or with “no labeled outcome” in the main causal analysis (see Main Methods-Disability Outcomes). For patients with no labeled outcome, any observed (i.e., actual) PDDS score was eligible for inclusion. For labeled patients, only PDDS scores occurring prior to target treatment (BCD or NTZ) initiation date were included in imputation model training. In cases of consecutive observations of PDDS scores in close proximity, we then excluded any PDDS scores that occurred within 1-month of a preceding score. Only data from patients with complete covariate data (and at least one observed PDDS score) were considered for the imputation analysis. We used the same set of pre-treatment covariates described in the main causal analysis as predictors in fitting these imputation models. In place of target treatment initiation date, predictor information was calculated using all data preceding the date of each PDDS observation. **eTables 1a-c** describe the imputation cohort derivation for the all-lookback, 12-month lookback, and 6-month lookback models, respectively.

We fit penalized, ordinal regression models, varying the following penalty and feature sets to optimize prediction performance. We compared all combinations of ridge versus LASSO penalty; 6-month, 12-month, and all-lookback periods; and the inclusion of all available versus knowledge graph-derived EHR features. We split data into training and test sets at patient level such that all PDDS observations of a given patient were either included as training or test data (i.e., no individual contributed to both the training and test data set). The training data set included 80% of available data, and the test data set included the remaining 20%. For each respective lookback period, the same training-test split was used across feature sets for fair comparison. Models were fit on the training set, and predictive performance and calibration assessed on the held-out test data set. Penalty parameters were tuned via 10-fold cross-validation and selected to minimize misclassification error across the 10 folds.

**eTable 2** summarizes the prediction performance of all fitted models, measured by two concordance metrics: concordance of predictions from each model with respect to the predicted class (class with the highest predicted probability) and with respect to the predicted PDDS risk (average of observed PDDS scores). We also present details on the cohort derivation and model calibration for the final model included to calculate covariates in the main causal analysis. The highest-performing PDDS imputation model (all-lookback ridge regression, using all available EHR features) was used to generate baseline PDDS risk as a covariate in the pre-treatment covariate set for causal analyses. The same model was also used to generate the 1-year post-treatment PDDS risk covariate (i.e., Year-1 PDDS Risk). **eFigure 1** summarizes the calibration of this model.

**Knowledge Graph-Derived Feature Selection**

The knowledge graph leverages the pre-trained embeddings of large-scale clinical concepts learned from summary level EHR data at two independent healthcare systems (~12,500,000 Veterans Health Administration patients and 60,000 Mass General Brigham Biobank patients) and the textual descriptions of the concepts embedded using CODER pre-trained language model.[1] The knowledge graph, through the Online Narrative and Codified feature Search Engine (ONCE) is publicly accessible via a web-application at https://shiny.parse-health.org/ONCE/, which provides further information on available codified and narrative electronic health record (EHR) features as well as instructions for use. For analyses in this study, feature sets for both codified and narrative EHR features were downloaded from the ONCE web application on November 24, 2023. We used the same knowledge graph-derived feature sets and the following process to pre-process data for both causal analysis and fitting PDDS imputation models.

Knowledge graph-derived features included specific codified (e.g., PheCode, CCS, RxNorm, LOINC) and narrative (i.e., CUI) features relevant to either MS or disability. We used the expanded set (see Clinical Study section in the ONCE web application) to search “multiple sclerosis” to identify features relevant to MS from the ONCE knowledge graph. We used the reduced set (see Phenotyping section in the ONCE web application) to search “disability” to identify features potentially relevant to disability.

To filter knowledge graph-derived codified and narrative EHR features (as described above), we first applied a pre-treatment feature frequency of at least 10% in the study population (or at least 5% in sensitivity analysis). Next, we removed any RxNorm codes or CUI features related to the target treatment classes of interest (i.e., BCD or NTZ). We also excluded PheCodes and CUIs specifically indicating MS and RxNorm codes related to prior standard-efficacy DMT use, as these features were separately included as standard covariates (see Main Methods-Confounders).

To summarize the feature selection pipeline, we identified clinically relevant features via the ONCE knowledge graph, removed rare features using >10% non-zero (or >5% for sensitivity analysis) frequency threshold, and removed BCD or NTZ related RxNorm codes and CUIs. A comprehensive list of knowledge graph-derived features (prior to the frequency filter and sparsity check) is detailed in **eTables 4a-4d**.

**Detailed Causal Analysis**

To use both EHR and registry data, we calculated the comparative treatment effect in a semi-supervised, doubly-robust IPW method weighted by the double-index propensity score (DiPS) as proposed by Cheng et al.[2,3] This method enables efficient, consistent estimation of average treatment effects in a semi-supervised setting. Briefly, the doubly-robust IPW treatment effect estimator differs from the traditional IPW estimator by using the DiPS as the inverse weight and the imputed outcomes in settings without labeled outcome observations. First, the DiPS weight is a kernel-smoothed propensity score estimate, which is a function of both an initial treatment assignment model and an initial outcome model. Second, the DiPS weight is further used as a covariate in a separate outcome imputation model to generate outcome labels. The resulting estimator is not only doubly robust (i.e., consistent when either the initial treatment or initial outcome model is correctly specified) but also consistent even when the outcome imputation model does not perfectly predict true outcome status. Moreover, the method allows covariate selection using penalized regression, specifically adaptive LASSO. Thus, another novelty of the method is effective adjustment of confounders selected from a high-dimensional covariate set from EHR in the treatment and outcome models that underlie the kernel-smoothed propensity score and the outcome imputation model. A high-level overview of the analysis is presented in the Main Figure 1.

The outcome imputation model allowed post-treatment, surrogate information associated with the true outcome status. We thus considered two sets of data, pre- and post-treatment. Both sets of data included the same confounders described in the Main Methods-Confounders section. Pre-treatment information included all observed data before target treatment initiation date. Post-treatment information included all pre-treatment information and any additional information up to 1-year following target treatment initiation date. The post-treatment data included three additional covariates: a binary indicator of target treatment membership (BCD or NTZ), an (inverse) DiPS weight of treatment group, and a measure of 1-year post-treatment disability risk. For patients with observed PDDS scores within 1-year post-treatment initiation, the 1-year disability covariate was simply the mean PDDS score. For patients with no observed PDDS scores during the 1-year post-treatment initiation period, we calculated the predicted mean-PDDS score for each patient, using the previously fit PDDS imputation models and post-treatment confounder data.

We began by calculating the DiPS estimator using pre-treatment information. This double-index propensity score was a kernel-smoothing estimator that used information from both initial treatment class and outcome regression models. We first fit adaptive LASSO regression for the initial treatment class and outcome models. We standardized all features and tuned the penalty parameter via 5-fold cross-validation. In settings with and without labeled outcome observations, we then calculated, normalized, and transformed the linear predictors from each model by the standard Gaussian distribution function. We then regressed treatment status on the resulting, transformed linear predictors using kernel smoothing to calculate the DiPS. Finally, we applied a trimming rule, truncating propensity score values to lie between 0.1 and 0.9 for numerical stability.[4]

For the imputation model of sustained change outcomes (i.e., worsening and improvement), we then fit an adaptive LASSO model using post-treatment covariate data, including the kernel-smoothed propensity score and the 1-year post-treatment disability risk (as described earlier). The penalty parameter was again tuned by 5-fold cross-validation. The imputation model was trained using only labeled outcome data. We then imputed sustained change outcome in settings without labeled outcome observations and used the observed sustained change outcome in settings with labeled observations. Finally, we calculated the treatment effect as the difference in mean outcome between the BCD and NTZ group, weighted by the DiPS of each observation.

The above process generated a point estimate for the treatment effect of interest. To generate confidence intervals of the estimate, we employed a perturbation resampling procedure. Specifically, we repeated the above process over 1,000 iterations, where each observation was given a random weight for each iteration. We generated the random weights from a distribution with mean and variance equal to one. The 2.5th and 97.5th quantile observations were then taken as the limits of the 95% confidence interval. We calculated p-values through a data-depth inversion procedure to translate confidence interval into tests of statistical significance.[5]

### Power Analysis

The power analysis was conducted under the additional assumption that our estimator of the average treatment effect (ATE) is asymptotically normal. As the asymptotic distribution and variance of the DiPS-based ATE estimator is not fully characterized,[2,3] we calculated the estimate of the standard error by taking the mean absolute deviation over the 1,000 perturbed estimates. We then used this as a variance estimate for a power analysis of testing the difference in normal means. We used the same perturbation scheme to estimate the standard error in the observed-case analysis (i.e., using labeled data only) as a comparator. Acknowledging that a completely post-hoc study of power analysis may be misleading[6], we mitigated a pitfall of the post-hoc power analyses by using only the standard error from our estimation procedure. Specifically, we used the estimated standard error to calculate the minimum detectable effect size, that is the minimum risk difference for which we have a pre-specified (i.e., 80%) power threshold to detect as statistically significant.

### eTables and eFigures

Pertinent supplementary tables and figures are included below. Additional supplementary results are available at this project's GitHub repository at <https://github.com/xialab2016/BCD_NTZ_SemiSupervisedCausal>.

### References

1. Yuan Z, Zhao Z, Sun H, Li J, Wang F, Yu S. CODER: Knowledge-infused cross-lingual medical term embedding for term normalization. *J Biomed Inform*. 2022;126:103983. doi:10.1016/j.jbi.2021.103983
2. Cheng D, Chakrabortty A, Ananthakrishnan AN, Cai T. Estimating average treatment effects with a double-index propensity score. *Biometrics*. 2020;76(3):767-777. doi:10.1111/biom.13195
3. Cheng D, Ananthakrishnan AN, Cai T. Robust and efficient semi-supervised estimation of average treatment effects with application to electronic health records data. *Biometrics*. 2021;77(2):413-423. doi:10.1111/biom.13298
4. Crump RK, Hotz VJ, Imbens GW, Mitnik OA. Dealing with Limited Overlap in Estimation of Average Treatment Effects. *Biometrika*. 2009;96(1):187-199.
5. Liu RY, Singh K. Notions of Limiting P Values Based on Data Depth and Bootstrap. *J Am Stat Assoc*. 1997;92(437):266-277. doi:10.1080/01621459.1997.10473624
6. Althouse AD. Post Hoc Power: Not Empowering, Just Misleading. *J Surg Res*. 2021;259:A3-A6. doi:10.1016/j.jss.2019.10.049

### eFigures

### eFigure 1. Calibration of a PDDS Imputation Model with an All-Lookback Period and All Available EHR Features

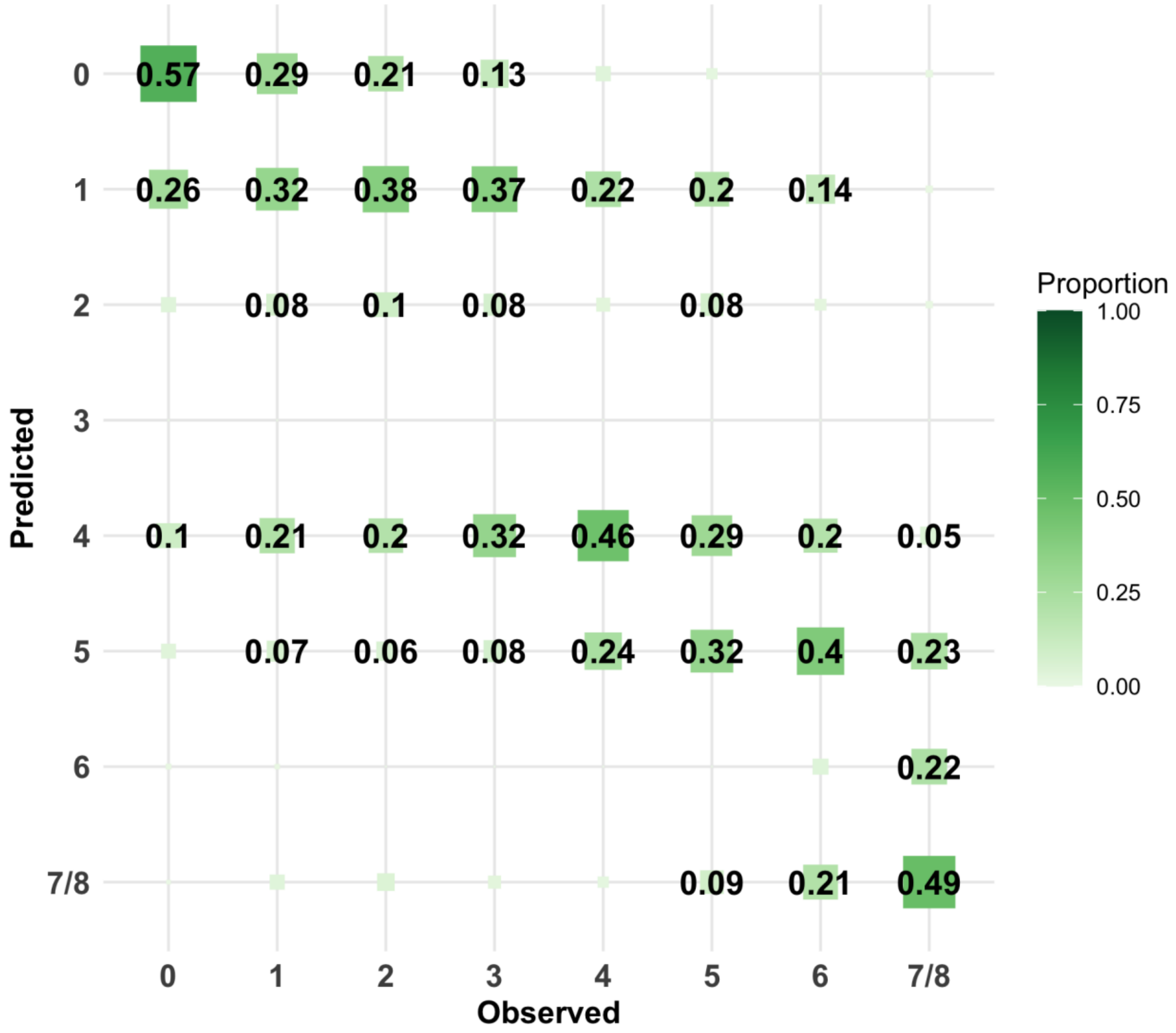

Numbers correspond to the column proportion of observations in each cell. For example, 0.57 in the top left indicates that among all observations where the true PDDS score was 0, we predicted 0 in 0.57 (or 57%) of the observations. Only values >0.05 (5%) reported. A perfect model would demonstrate values of 1 across the top-left to bottom-right diagonal.

**Abbreviations:** EHR, Electronic Healthcare Record. PDDS, Patient Determined Disease Steps.

### eFigure 2. Average Treatment Effects for 2- and 3-year Evaluation Windows

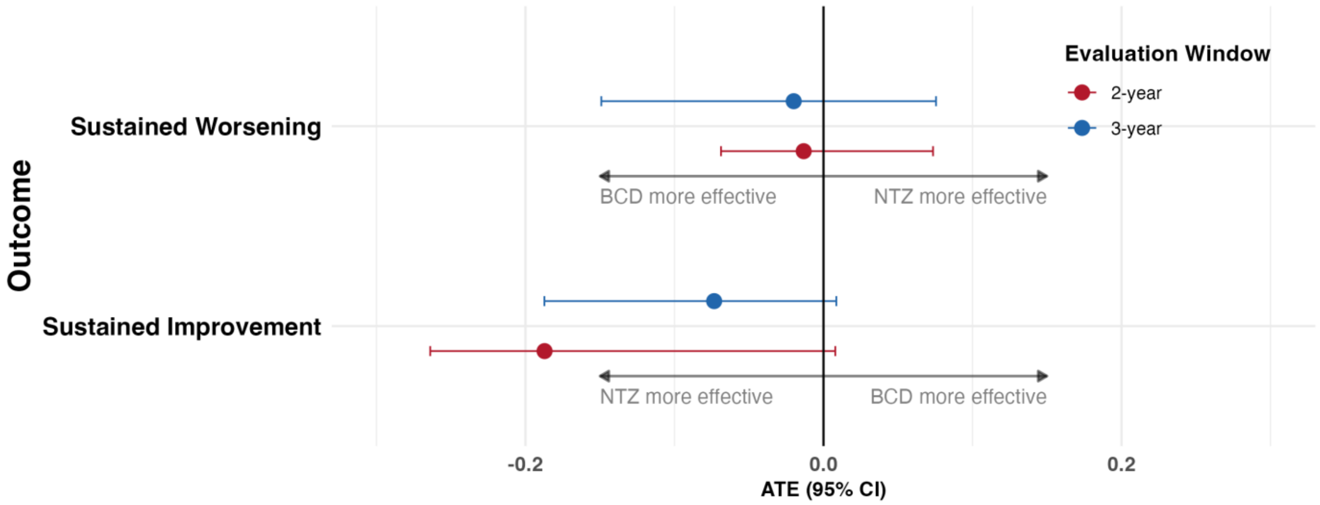

Points correspond to estimates of average treatment effect (ATE). Lines correspond to 95% confidence intervals estimated by perturbation analysis. With NTZ as the reference treatment class, a positive ATE for the sustained worsening outcome would indicate BCD as less effective than NTZ in reducing patient-reported disability worsening. Conversely, a positive ATE for the sustained improvement outcome would indicate BCD as more effective than NTZ in promoting patient-reported disability improvement.
**Abbreviations**: ATE, Average Treatment Effect. BCD, B-Cell Depletion. CI, Confidence Interval. NTZ, Natalizumab.

### eFigure 3a. Sensitivity Results for Sustained Worsening Models

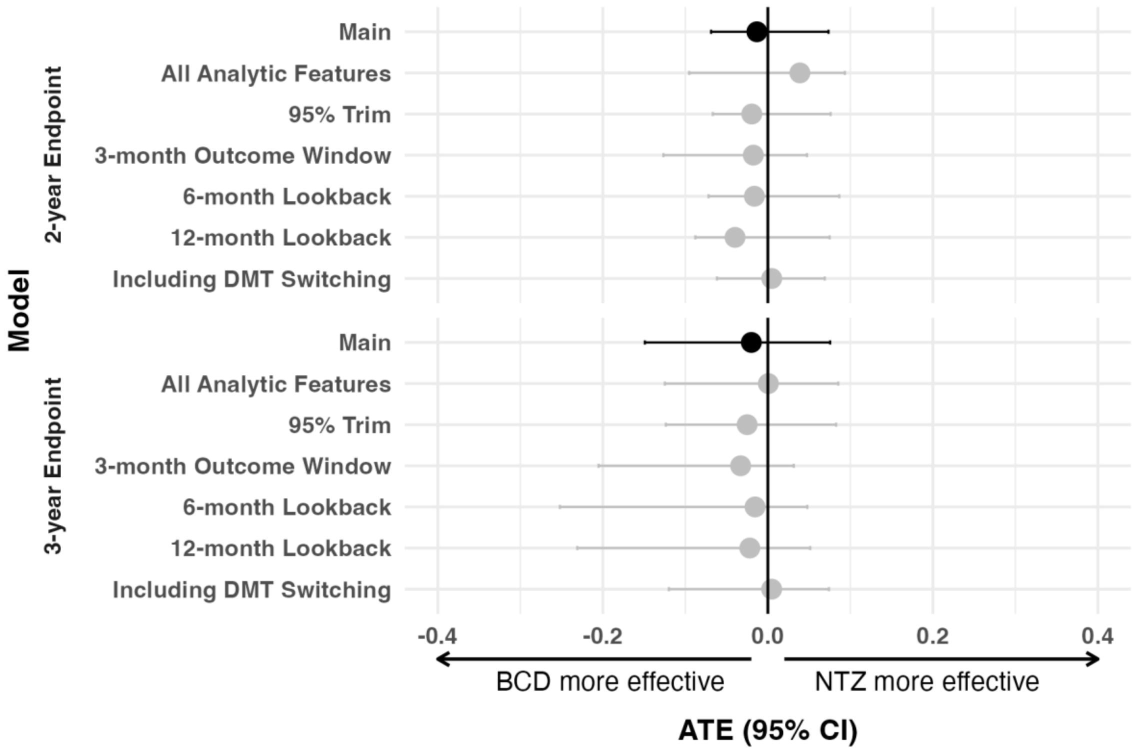

Points correspond to estimates of average treatment effect (ATE). Lines correspond to 95% confidence intervals estimated by perturbation analysis. Results are grouped by end point (2- or 3-years after treatment initiation). Each row corresponds to the result of a respective sensitivity analysis for the sustained worsening outcome. With NTZ as the reference treatment class, a positive ATE for the sustained worsening outcome would indicate BCD as less effective than NTZ in reducing patient-reported disability worsening.
**Abbreviations**: ATE, Average Treatment Effect. BCD, B-Cell Depletion. CI, Confidence Interval. DMT, Disease-Modifying Therapy. NTZ, Natalizumab.

### eFigure 3b. Sensitivity Results for Sustained Improvement Models

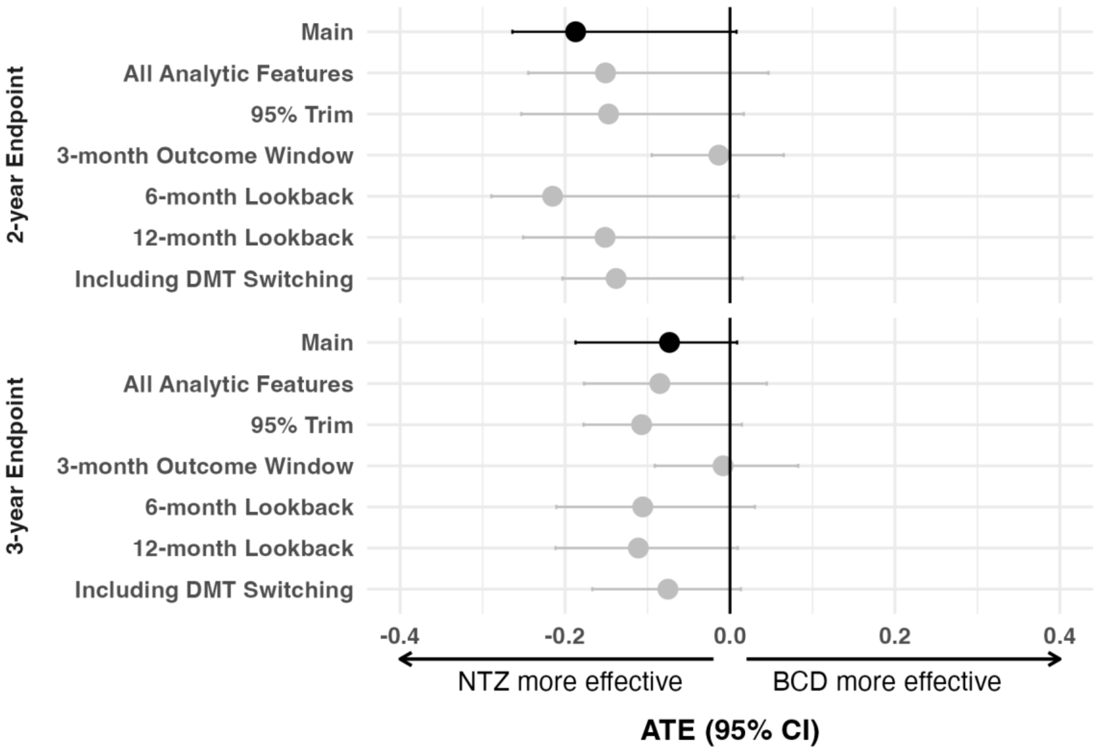

Points correspond to estimates of average treatment effect (ATE). Lines correspond to 95% confidence intervals estimated by perturbation analysis. Results are grouped by end point (2- or 3-years after treatment initiation). Each row corresponds to the result of a respective sensitivity analysis for the sustained improvement outcome. A positive ATE for the sustained improvement outcome would indicate BCD as more effective than NTZ in promoting patient-reported disability improvement.

**Abbreviations**: ATE, Average Treatment Effect. BCD, B-Cell Depletion. CI, Confidence Interval. DMT, Disease-Modifying Therapy. NTZ, Natalizumab.

### eTables

#### eTable 1a. Cohort derivation for All-Lookback Cohorts in PDDS Imputation Modeling

| Step | Patients (N) | PDDS Observations (n) |
|---|---|---|
| (0) Observations of any PDDS Score | 1,985 | 17,626 |
| (1) PDDS Scores Occurring 1+ Months Apart | 1,985 | 14,294 |
| (2) Patients with Complete Demographic and Clinical Covariates | 1,680 | 13,258 |
| (4) Observations Unused in Causal Analysis to Categorize Post-treatment Sustained-Change Outcome | 1,596 | 10,658 |

**Abbreviations:** PDDS, Patient Determined Disease Steps.

#### eTable 1b. Cohort Derivation for 12-month Lookback Cohorts in PDDS Imputation Modeling

| Step | Patients (N) | PDDS Observations (n) |
|---|---|---|
| (0) Observations of any PDDS Score | 1,985 | 17,626 |
| (1) PDDS Scores Occurring 1+ Months Apart | 1,985 | 14,294 |
| (2) Patients with Complete Demographic & Clinical Covariates | 1,676 | 13,164 |
| (4) Observations Unused in Causal Analysis to Categorize Post-treatment Sustained-Change Outcome | 1,592 | 10,572 |

**Abbreviations:** PDDS, Patient Determined Disease Steps.

#### eTable 1c. Cohort Derivation for 6-month Lookback Cohorts in PDDS Imputation Modeling

| Step | Patients (N) | PDDS Observations (n) |
|---|---|---|
| (0) Observations of any PDDS Score | 1,985 | 17,626 |
| (1) PDDS Scores Occurring 1+ Months Apart | 1,985 | 14,294 |
| (2) Patients with Complete Demographic & Clinical Covariates | 1,669 | 13,027 |
| (4) Observations Unused in Causal Analysis to Categorize Post-treatment Sustained-Change Outcome | 1,585 | 10,456 |

**Abbreviations:** PDDS, Patient Determined Disease Steps.

### eTable 2. Summary of PDDS Imputation Model Performance by Penalty, Covariate Set, and Lookback Window

| Lookback | Penalty | Covariates | Used in Causal Analysis | Concordance (PDDS Category) | Concordance (Mean PDDS) |
|---|---|---|---|---|---|
| **Inf months** | **ridge** | **All Available Features** | **Yes** | **0.783** | **0.807** |
| 12 months | ridge | All Available Features | No | 0.770 | 0.804 |
| Inf months | ridge | Knowledge Graph Derived | No | 0.764 | 0.803 |
| 6 months | ridge | All Available Features | No | 0.754 | 0.800 |
| 12 months | ridge | Knowledge Graph Derived | No | 0.725 | 0.799 |
| Inf months | lasso | Knowledge Graph Derived | No | 0.768 | 0.799 |
| 12 months | lasso | Knowledge Graph Derived | No | 0.763 | 0.798 |
| 12 months | lasso | All Available Features | No | 0.758 | 0.789 |
| 6 months | lasso | All Available Features | No | 0.764 | 0.786 |
| 6 months | ridge | Knowledge Graph Derived | No | 0.717 | 0.782 |
| Inf months | lasso | All Available Features | No | 0.767 | 0.781 |
| 6 months | lasso | Knowledge Graph Derived | No | 0.744 | 0.778 |

"All available features" correspond to inclusion of any available EHR feature (PheCode, CCS, RxNorm, LOINC) observed in at least 10% of patients (and included after applying the PheCode roll-up procedure). Concordance was calculated using the predicted PDDS risk, assigned as the PDDS category with highest probability of membership. Categories 7 & 8 (represented highest level of disability) were combined due to sparsity of membership in category 8 (<1% of observations).

**Abbreviations:** PDDS, Patient Determined Disease Steps.

### eTable 3a. Knowledge Graph-Derived, Codified EHR Features Related to Multiple Sclerosis

| Variable | Description |
| --- | --- |
| RXNORM:897018 | dalfampridine |
| PheCode:596.5 | functional disorders of bladder |
| PheCode:341 | other demyelinating diseases of central nervous system |
| RXNORM:30125 | modafinil |
| PheCode:296 | mood disorders |
| PheCode:300 | anxiety disorders |
| RXNORM:1292 | baclofen |
| PheCode:296.2 | depression |
| PheCode:377.3 | optic neuritis/neuropathy |
| PheCode:334 | degenerative disease of the spinal cord |
| PheCode:350.1 | abnormal involuntary movements |
| PheCode:296.22 | major depressive disorder |
| PheCode:377.1 | optic atrophy |
| PheCode:323.2 | acute (transverse) myelitis |
| PheCode:707.1 | decubitus ulcer |
| PheCode:596 | other disorders of bladder |
| PheCode:709.3 | systemic sclerosis |
| RXNORM:6832 | methenamine |
| PheCode:300.9 | posttraumatic stress disorder |
| PheCode:290.16 | vascular dementia |
| PheCode:300.1 | anxiety disorder |
| PheCode:798 | malaise and fatigue |

### eTable 3b. Knowledge Graph-Derived, Narrative EHR Features Related to Multiple Sclerosis

| CUI | Variable |
|---|---|
| C0026769 | Multiple Sclerosis |
| C0751967 | Multiple Sclerosis, Relapsing-Remitting |
| C0011304 | Demyelination |
| C0029134 | Optic Neuritis |
| C0011303 | Demyelinating Diseases |
| C0011570 | Mental Depression |
| C0231170 | Disability |
| C0069426 | Oligoclonal Bands (protein) |
| C2752009 | White matter lesion |
| C1579931 | Depressed - symptom |
| C0035020 | Relapse |
| C0026838 | Muscle Spasticity |
| C0011581 | Depressive disorder |
| C0004093 | Asthenia |
| C0682148 | Disability status |
| C0087136 | Unmarried |
| C0423551 | Sensory symptoms |
| C3540014 | CENTRAL NERVOUS SYSTEM DIAGNOSTIC RADIOPHARMACEUTICALS |
| C0163712 | Relate - vinyl resin |
| C0202205 | Oligoclonal protein measurement |
| C0004268 | Attention |
| C0066677 | modafinil |
| C0344315 | Depressed mood |
| C5203119 | Intensity and Distress 5 |
| C0010957 | Tissue damage |
| C1558916 | Adverse Event Associated with Syndromes |

| CUI | Variable |
|---|---|
| C0878575 | Peripheral demyelination |
| C1457887 | Symptoms |
| C5202921 | RECIL PD |
| C0241224 | Spinal cord lesion |
| C5202991 | IMWG Progressive Disease |
| C0017639 | Gliosis |
| C0015672 | Fatigue |
| C3539781 | Progressive cGVHD |
| C2004461 | Bowel dysfunction |
| C3714552 | Weakness |
| C4551520 | Intention tremor |
| C0338656 | Impaired cognition |
| C1963758 | Immunomodulation |
| C0339662 | Afferent Pupillary Defect |
| C4050309 | Central Nervous System Involvement |
| C0040997 | Trigeminal Neuralgia |
| C0036429 | Sclerosis |
| C0235946 | Cerebral atrophy |
| C0024485 | Magnetic Resonance Imaging |
| C3665386 | Abnormal vision |
| C0237607 | Practice Experience |
| C4723839 | irPD (Immune-Related Response Criteria) |
| C0004609 | Baclofen |
| C5202924 | Global Progressive Disease in Skin |
| C0741548 | bladder symptoms |
| C0037763 | Spasm |
| C0221423 | Illness (finding) |

| CUI | Variable |
|---|---|
| C1522240 | Process |
| C0221198 | Lesion |
| C4084203 | Improved - answer to question |
| C3887651 | Palsy |
| C0039082 | Syndrome |
| C0004134 | Ataxia |
| C1561270 | Adverse Event Associated with Neurology |
| C0436596 | On examination - apathetic |
| C1299586 | Has difficulty doing (qualifier value) |
| C0043143 | wheelchair |
| C0443306 | Spastic |
| C0027627 | Neoplasm Metastasis |
| C0025260 | Memory |
| C0525041 | Neurobehavioral Manifestations |
| C4263551 | Multisection for pediatrics |
| C1269683 | Major Depressive Disorder |
| C4721418 | Legally Separated |
| C0232841 | Bladder dysfunction |
| C0012569 | Diplopia |
| C0242656 | Disease Progression |
| C3263723 | Traumatic injury |
| C4042908 | Seroconversion |
| C0042024 | Urinary Incontinence |
| C0849867 | Generalized illness |
| C0004364 | Autoimmune Diseases |
| C2239268 | magnets - physical therapy modality |
| C0013362 | Dysarthria |

| **CUI** | **Variable** |
|---|---|
| C0002403 | Amantadine |
| C0422943 | Visual symptoms |
| C5202689 | HIV Seroconversion |
| C0021368 | Inflammation |
| C0235146 | Euphoric mood |
| C4551583 | Cerebral cortical atrophy |
| C0003469 | Anxiety Disorders |
| C0037928 | Spinal Cord Diseases |
| C5401372 | CTRP Disease Finding |
| C1291764 | Immune System Finding |
| C0025815 | Methylprednisolone |
| C0007758 | Cerebellar Ataxia |
| C0579152 | Bladder problem |
| C3665346 | Unspecified visual loss |
| C0012634 | Disease |
| C0018681 | Headache |
| C0041696 | Unipolar Depression |
| C3539106 | Sufficiently defined concept definition status (core metadata concept) |
| C0700327 | Memory observations |
| C0684336 | Impaired health |
| C0234518 | Slurred speech |
| C0004083 | Mental association |
| C0005956 | Bone Marrow Diseases |
| C0271051 | Macular retinal edema |
| C0036454 | Scotoma |
| C0086045 | Mental concentration |
| C0025611 | Methamphetamine |

| CUI | Variable |
|---|---|
| C1556682 | Adverse Event Associated with Infection |
| C1363945 | Therapy Object (animal model) |
| C0596545 | Experience |
| C0270612 | Leukoencephalopathy |
| C3263722 | Traumatic AND/OR non-traumatic injury |
| C0003537 | Aphasia |
| C0525045 | Mood Disorders |
| C0042571 | Vertigo |
| C0019202 | Hepatolenticular Degeneration |
| C0231217 | Multiple symptoms |
| C0544452 | Disease remission |
| C0038435 | Stress |
| C0439044 | Living Alone |
| C0028643 | Numbness |
| C0003467 | Anxiety |
| C0235031 | Neurologic Symptoms |
| C0009450 | Communicable Diseases |
| C0036572 | Seizures |
| C0332461 | Plaque (lesion) |
| C0009024 | Clonus |
| C0022423 | Judgment |
| C3839460 | Nonprogressive |
| C0549622 | Sexual Dysfunction |
| C5142828 | Full recovery |
| C0040822 | Tremor |
| C0543488 | Interested |
| C0085633 | Mood swings |

| **CUI** | **Variable** |
|---|---|
| C0021345 | Infectious Mononucleosis |
| C0030554 | Paresthesia |
| C0000768 | Congenital Abnormality |
| C0004368 | Autoimmune state |
| C0679575 | Neuroimaging |
| C4028269 | Nuclear magnetic resonance imaging brain |
| C1306577 | Death (finding) |
| C1509143 | Physical assessment findings |
| C0005525 | Biological Response Modifiers |
| C0522224 | Paralysed |
| C2936842 | Vitamin D [EPC] |
| C1964257 | Observation - diagnostic procedure |
| C0687702 | Cancer Remission |
| C1838681 | Rapidly progressive |
| C0014544 | Epilepsy |
| C0020580 | Hypesthesia |
| C0860603 | Anxiety symptoms |
| C0027404 | Narcolepsy |
| C0079595 | Imaging Techniques |
| C0456909 | Blindness |
| C3887506 | Hyperkinesia |
| C0558058 | Reflecting |
| C0229992 | Psyche structure |
| C0013781 | Shock from electric current |
| C0015967 | Fever |
| C4551761 | Excessive daytime sleepiness |
| C0277785 | Functional disorder |

| CUI | Variable |
|---|---|
| C0032989 | Multiple Pregnancy |
| C0005586 | Bipolar Disorder |
| C0008300 | Choice Behavior |
| C0009806 | Constipation |
| C0080274 | Urinary Retention |
| C0919758 | Vitamin D measurement |
| C0233496 | Aversion (finding) |
| C0042866 | Vitamin D |

Variable names of the concept unique identifiers were exported directly from the Unified Medical Language System (UMLS) metathesaurus browser definitions.

### eTable 3c. Knowledge Graph-Derived, Codified EHR Features Related to Disability

| Variable | Description |
|---|---|
| PheCode:296.2 | depression |
| PheCode:296.22 | major depressive disorder |
| PheCode:296 | mood disorders |
| PheCode:300.1 | anxiety disorder |
| PheCode:300 | anxiety disorders |
| CCS:218 | psychological and psychiatric evaluation and therapy |
| PheCode:300.4 | dysthymic disorder |
| PheCode:300.9 | posttraumatic stress disorder |
| PheCode:317.1 | alcoholism |
| PheCode:301 | personality disorders |
| PheCode:297.1 | suicidal ideation |
| PheCode:317 | alcohol-related disorders |
| PheCode:300.11 | generalized anxiety disorder |
| PheCode:316 | substance addiction and disorders |
| PheCode:300.13 | phobia |
| PheCode:304 | adjustment reaction |
| PheCode:295.3 | psychosis |
| PheCode:327.4 | insomnia |
| PheCode:295 | schizophrenia and other psychotic disorders |
| PheCode:300.12 | agorophobia, social phobia, and panic disorder |
| PheCode:315.3 | mental retardation |
| PheCode:969 | poisoning by psychotropic agents |
| PheCode:301.2 | antisocial/borderline personality disorder |
| PheCode:300.8 | acute reaction to stress |
| PheCode:312 | conduct disorders |
| PheCode:338 | pain |

| Variable | Description |
|---|---|
| PheCode:760 | back pain |
| PheCode:313.3 | autism |
| PheCode:318 | tobacco use disorder |
| PheCode:340 | migraine |
| PheCode:295.2 | paranoid disorders |
| PheCode:296.1 | bipolar |
| PheCode:295.1 | schizophrenia |
| PheCode:305.21 | anorexia nervosa |
| PheCode:313.1 | attention deficit hyperactivity disorder |
| PheCode:773 | pain in limb |
| LOINC:3349-8 | loinc:amphetamines |
| LOINC:3397-7 | loinc:cocaine |
| PheCode:303.1 | dissociative disorder |
| PheCode:745 | pain in joint |
| PheCode:327 | sleep disorders |
| CCS:237 | ancillary services |
| PheCode:303.3 | psychogenic disorder |
| LOINC:3879-4 | loinc:opiates |
| LOINC:18282-4 | loinc:cannabinoids |
| PheCode:300.3 | obsessive-compulsive disorders |
| LOINC:3390-2 | loinc:benzodiazepines |
| PheCode:305.2 | eating disorder |
| PheCode:297.2 | suicide or self-inflicted injury |
| PheCode:963 | poisoning by primarily systemic agents |
| PheCode:338.2 | chronic pain |
| PheCode:789 | nausea and vomiting |
| LOINC:14314-9 | cocaine, urine (group:urcoca) |

| Variable | Description |
|---|---|
| LOINC:17384-9 | opiates, urine (group:uopi) |
| PheCode:292.6 | hallucinations |
| PheCode:278.1 | obesity |
| LOINC:16369-1 | amphetamine(s), urine (group:uamph) |
| PheCode:761 | cervicalgia |
| RXNORM:42347 | bupropion |
| PheCode:290.13 | senile dementia |
| PheCode:306 | other mental disorder |
| PheCode:292.3 | memory loss |
| PheCode:530.11 | gerd |
| LOINC:14316-4 | benzodiazepines, urine (group:ubenz) |
| PheCode:306.9 | tension headache |
| PheCode:704.1 | alopecia |
| PheCode:737.2 | lordosis (acquired) |
| LOINC:3773-9 | loinc:methadone |
| PheCode:313 | pervasive developmental disorders |
| PheCode:315.1 | learning disorder |
| PheCode:563 | constipation |
| PheCode:290.1 | dementias |
| PheCode:260.6 | anorexia |
| PheCode:312.3 | impulse control disorder |
| PheCode:303.4 | somatoform disorder |
| LOINC:16254-5 | phencyclidine, urine (group:urpcp) |
| LOINC:14624-1 | barbiturates, urine (group:ubarb) |
| PheCode:704.2 | hirsutism |
| PheCode:599.4 | urinary incontinence |
| LOINC:X7003-7 | ethanol (tox panel) (group:tox1) |

| Variable | Description |
|---|---|
| PheCode:798 | malaise and fatigue |
| PheCode:244 | hypothyroidism |
| PheCode:293 | symptoms involving head and neck |
| PheCode:264 | lack of normal physiological development |
| PheCode:355.1 | chronic pain syndrome |
| LOINC:14313-1 | thc/cannabinoids, urine (group:uthc) |
| PheCode:389.4 | tinnitus |
| PheCode:303 | psychogenic and somatoform disorders |
| LOINC:5643-2 | loinc:ethanol |
| PheCode:327.3 | sleep apnea |
| LOINC:35635-2 | trazodone (group:trazo) |
| PheCode:799 | debility unspecified |
| PheCode:605 | erectile dysfunction [ed] |
| RXNORM:1547099 | suvorexant |
| RXNORM:3554 | disulfiram |
| LOINC:X7011-0 | other findings (tox) (group:otfi) |
| LOINC:3494-2 | clonazepam (group:clonaz) |
| PheCode:339 | other headache syndromes |
| LOINC:10998-3 | loinc:oxycodone |
| PheCode:278.4 | abnormal weight gain |
| PheCode:536.3 | gastroparesis |
| RXNORM:72625 | duloxetine |
| PheCode:389 | hearing loss |
| PheCode:342 | hemiplegia |
| RXNORM:4493 | fluoxetine |
| PheCode:301.1 | schizoid personality disorder |
| PheCode:785 | abdominal pain |

| Variable | Description |
|---|---|
| PheCode:755.4 | congenital anomalies of upper limb, including shoulder girdle |
| PheCode:292.4 | altered mental status |
| RXNORM:5553 | hydroxyzine |
| LOINC:13572-3 | nordiazepam (group:ddiaz) |
| PheCode:1005 | other symptoms |
| LOINC:5644-0 | loinc:ethanol |
| RXNORM:10737 | trazodone |
| PheCode:975 | poisoning by agents primarily acting on the smooth and skeletal muscles and respiratory system |
| PheCode:367.1 | myopia |
| PheCode:625.1 | dyspareunia |
| PheCode:261.1 | vitamin a deficiency |
| PheCode:966 | poisoning by anticonvulsants and anti-parkinsonism drugs |
| PheCode:292.12 | symbolic dysfunction |
| RXNORM:3638 | doxepin |
| LOINC:X7004-5 | ats (tox panel) (group:tox2) |
| PheCode:350.3 | lack of coordination |
| PheCode:599.3 | Dysuria |
| PheCode:1010.7 | persons with potential health hazards related to socioeconomic, psychosocial, and other circumstances |
| RXNORM:1455099 | vortioxetine |
| LOINC:X7005-2 | barbiturates (tox panel) (group:tox3) |
| CCS:219 | alcohol and drug rehabilitation/detoxification |
| PheCode:626.2 | dysmenorrhea |
| LOINC:2106-3 | loinc:choriogonadotropin (pregnancy test) |
| PheCode:710.2 | periostitis |

### eTable 3d. Knowledge Graph-Derived, Narrative EHR Features Related to Disability

| CUI | Variable |
|---|---|
| C0231170 | Disability |
| C3714756 | Intellectual Disability |
| C0684336 | Impaired health |
| C5203119 | Intensity and Distress 5 |
| C0008073 | Developmental Disabilities |
| C4084203 | Improved - answer to question |
| C0595998 | Household composition |
| C0751265 | Learning Disabilities |
| C0004936 | Mental disorders |
| C1299586 | Has difficulty doing (qualifier value) |
| C0163712 | Relate - vinyl resin |
| C0229992 | Psyche structure |
| C0596545 | Experience |
| C1509143 | Physical assessment findings |
| C0087136 | Unmarried |
| C0004268 | Attention |
| C0237607 | Practice Experience |
| C0004271 | Attitude |
| C3665347 | Visual Impairment |
| C0004083 | Mental association |
| C0231172 | handicapping condition |
| C0700327 | Memory observations |
| C0025260 | Memory |
| C0543488 | Interested |
| C0221423 | Illness (finding) |
| C0004448 | Awareness |

| CUI | Variable |
|---|---|
| C0557351 | Employed |
| C0041674 | Unemployment |
| C0678856 | skill |
| C0011570 | Mental Depression |
| C2239122 | Social history of activities |
| C0023185 | Learning |
| C0162429 | Malnutrition |
| C0597198 | Performance |
| C0025362 | Mental Retardation |
| C0013658 | Educational Status |
| C0455498 | History of - psychiatric disorder |
| C0456909 | Blindness |
| C1306597 | Psychiatric problem |
| C1522240 | Process |
| C1579931 | Depressed - symptom |
| C0030971 | Perception |
| C0476254 | Dyslexia |
| C3263723 | Traumatic injury |
| C0010957 | Tissue damage |
| C0025611 | Methamphetamine |
| C0542559 | contextual factors |
| C0001721 | Affect (mental function) |
| C0424605 | Developmental delay (disorder) |
| C0034991 | Rehabilitation therapy |
| C3495449 | Mobility aid |
| C0043143 | wheelchair |
| C1384666 | hearing impairment |
| C0002957 | Anger |

| CUI | Variable |
|---|---|
| C0007237 | Encounter due to care involving use of rehabilitation procedures |
| C0011581 | Depressive disorder |
| C0311392 | Physical findings |
| C1457887 | Symptoms |
| C0018772 | Hearing Loss, Partial |
| C0205082 | Severe (severity modifier) |
| C0004352 | Autistic Disorder |
| C0558058 | Reflecting |
| C0038272 | Stereotyping |
| C5399832 | Inclusion Body (finding) |
| C1550518 | incapable |
| C0007952 | Personality Character |
| C4551887 | birth (history) |
| C1444648 | Offered |
| C1171285 | Enabling |
| C0332218 | Difficult (qualifier value) |
| C0277787 | Social stigmata |
| C0344315 | Depressed mood |
| C0680095 | Personal failure |
| C0033975 | Psychotic Disorders |
| C0441722 | Force |
| C0012634 | Disease |
| C4745084 | Medical Condition |
| C0876926 | Traumatic Brain Injury |
| C0233820 | Insight |
| C5201148 | Moderate |
| C3263722 | Traumatic AND/OR non-traumatic injury |
| C0185117 | Expression procedure |

| CUI | Variable |
|---|---|
| C0023186 | Academic skill disorder |
| C1548428 | Referral type - Psychiatric |
| C0184511 | Improved |
| C0349588 | Short stature |
| C0425245 | Mobility as a finding |
| C0557874 | Global developmental delay |
| C0162340 | Comprehension |
| C0557061 | Discussion (procedure) |
| C0600138 | Does play |
| C0026769 | Multiple Sclerosis |
| C0332840 | Amputated structure (morphologic abnormality) |
| C0178499 | Base |
| C0022423 | Judgment |
| C0439044 | Living Alone |
| C0013987 | Emotions |
| C1306577 | Death (finding) |
| C0765629 | President brand of dental material |
| C0424939 | Learning difficulties |
| C2733607 | Developmentally disabled (finding) |
| C1527305 | Feelings |
| C0013146 | Drug abuse |
| C0036341 | Schizophrenia |
| C0013336 | Dwarfism |
| C0011053 | Deafness |
| C3526598 | Psychiatric service |
| C0005586 | Bipolar Disorder |
| C0038436 | Post-Traumatic Stress Disorder |
| C0562342 | Empowered |

| CUI | Variable |
|---|---|
| C1559081 | Adverse Event Associated with Death |
| C0522224 | Paralysed |
| C0338656 | Impaired cognition |
| C0000768 | Congenital Abnormality |
| C0001807 | Aggressive behavior |
| C0013080 | Down Syndrome |
| C0848067 | Mental problem |
| C0036597 | Self Esteem |
| C2237041 | SHOX gene with short stature |
| C0036605 | Self-Help Devices |
| C0162425 | Intention - mental process |
| C0009671 | Conflict (Psychology) |
| C3714660 | Trauma |
| C1273518 | Responsible to |
| C5441521 | Complaint (finding) |
| C0024841 | Marriage, life event |
| C0231303 | Distress |
| C2347509 | Physical Shift |
| C3887873 | Hearing Loss |
| C0600116 | Does speak |
| C1263846 | Attention deficit hyperactivity disorder |
| C3539106 | Sufficiently defined concept definition status (core metadata concept) |
| C4759845 | CTCAE v4 Grade 2 |
| C0000924 | Accidents |
| C2029884 | hearing loss by exam |
| C1457898 | Growth & development aspects |
| C3844700 | Two or more |

| CUI | Variable |
|---|---|
| C0037420 | Social Interaction |
| C0424215 | Sense of identity (observable entity) |
| C0042798 | Low Vision |
| C0683525 | treatment options |
| C0018674 | Craniocerebral Trauma |
| C0018786 | Hearing Tests |
| C0009241 | Cognition Disorders |
| C1269683 | Major Depressive Disorder |
| C1704241 | complex (molecular entity) |
| C0233514 | Abnormal behavior |
| C0566001 | Does communicate |
| C0014544 | Epilepsy |
| C0678341 | Responsibility |
| C5452990 | Helped |
| C2937289 | Adapt (substance) |
| C0459920 | Abstract thinking ability |
| C0424595 | Undifferentiated illness |
| C0003469 | Anxiety Disorders |
| C0392673 | Adaptation |
| C0679199 | Strategy |
| C0425382 | Personal status - Adopted |
| C0041696 | Unipolar Depression |
| C3714578 | Fix |
| C0681405 | Preschool Completion |
| C1821973 | Vulnerability |
| C0237529 | Self Confidence |
| C4316940 | Joy |

| CUI | Variable |
|---|---|
| C2220266 | exposure history |
| C0424589 | Vitality |
| C0025353 | mental health |
| C1444656 | Indicated |
| C3275042 | Abandoned Lead |
| C0039082 | Syndrome |
| C2745965 | Emergencies [Disease/Finding] |
| C2242890 | History of deprivation |
| C3540014 | CENTRAL NERVOUS SYSTEM DIAGNOSTIC RADIOPHARMACEUTICALS |
| C0015726 | Fear (Mental Process) |
| C0027627 | Neoplasm Metastasis |
| C3714552 | Weakness |
| C1171947 | Commit Lozenge |
| C0441516 | Demand (clinical) |
| C2242855 | personal hygiene (history) |
| C0013956 | Emergency Situation |
| C0234856 | Speaking (activity) |
| C0740858 | Substance abuse problem |
| C0871633 | desire |
| C5234924 | Positive Attitude |
| C0332447 | Morphologically abnormal structure (morphologic abnormality) |
| C0449416 | Source |
| C0043251 | Wounds and Injuries |
| C0033211 | Problem Solving (mental process) |
| C1458132 | Treatment/Psychosocial Effects |
| C3887651 | Palsy |
| C0525045 | Mood Disorders |

| CUI | Variable |
|---|---|
| C0018379 | Guilt |
| C0278061 | Abnormal mental state |
| C0518609 | Consideration |
| C0270611 | Brain Injuries |
| C1559524 | Adverse Event Associated with Growth And Development |
| C0002688 | Amputation |
| C0871215 | Reading Disabilities |
| C0277814 | Sitting position |
| C1821293 | Worthlessness |
| C0020039 | Hostility |
| C0150055 | Chronic pain |
| C0425229 | Overcrowded in house |
| C0521874 | Victim of neglect (finding) |
| C1363945 | Therapy Object (animal model) |
| C0358514 | Diagnostic agents |
| C0039869 | Thinking, function |
| C0013126 | Intrinsic drive |
| C2169640 | recently raped (history) |
| C0022107 | Irritable Mood |
| C0028768 | Obsessive-Compulsive Disorder |
| C0274281 | Injury due to exposure to external cause |
| C1882365 | Phenomenon |
| C3887804 | Feeling upset |
| C2986546 | Target Lesion Identification |
| C0683323 | physical illness |
| C4738113 | Fatalities |
| C0026838 | Muscle Spasticity |
| C0679105 | pleasurable emotion |

| CUI | Variable |
|---|---|
| C0037937 | Spinal Injuries |
| C4263342 | Multisection metabolic |
| C0442797 | Decreasing |
| C4760315 | Obsessive compulsive disorder drugs |
| C2828386 | Pass (indicator) |
| C0424318 | Bullying |
| C0038435 | Stress |
| C0282350 | Sexual abuse |
| C0231173 | Invalidism |
| C0003467 | Anxiety |
| C0004930 | Behavior Disorders |
| C0018524 | Hallucinations |
| C0008679 | Chronic disease |
| C0231441 | Immobile |
| C0004044 | Asphyxia |
| C0860603 | Anxiety symptoms |
| C0020580 | Hypesthesia |
| C0035345 | Retirement |
| C0679006 | Decision |
| C5236074 | Identified By |
| C0009676 | Confusion |
| C0205161 | Abnormal |
| C4263551 | Multisection for pediatrics |
| C2700617 | Irritation - emotion |
| C0002658 | Amphetamine |
| C2186378 | Reported history of chronic illness |
| C0600109 | Willing |
| C0231337 | Senility |

| CUI | Variable |
| --- | --- |
| C0497327 | Dementia |

Variable names of the concept unique identifiers were exported directly from the Unified Medical Language System (UMLS) metathesaurus browser definitions.

### eTable 4a. Cohort Characteristics (by 2-Year Evaluation Window)

| Variable | No Sustained Change (n=114) | Sustained Improvement (n=44) | Sustained Worsening (n=59) | Unlabeled (n=1521) |
|---|---|---|---|---|
| Age at Target Treatment Initiation: years, mean (SD) | 41.51 (12.16) | 42.41 (10.56) | 46.36 (13.25) | 46.40 (13.07) |
| Self-Reported Gender: n (%) | | | | |
| Men | 31 (27.19%) | 6 (13.64%) | 22 (37.29%) | 429 (28.21%) |
| Women | 83 (72.81%) | 38 (86.36%) | 37 (62.71%) | 1,092 (71.79%) |
| Race and ethnicity: n (%) | | | | |
| White, Non-Hispanic | 97 (85.09%) | 40 (90.91%) | 55 (93.22%) | 1,315 (86.46%) |
| Other race and ethnic groups | 17 (14.91%) | 4 (9.09%) | 4 (6.78%) | 206 (13.54%) |
| Disease Duration: years, mean (SD) | 4.23 (5.16) | 3.76 (4.46) | 5.03 (5.31) | 5.39 (4.71) |
| Follow-up Duration: years, mean (SD) | 9.77 (5.13) | 10.12 (4.47) | 9.60 (4.99) | 9.25 (4.99) |
| Target Treatment Class: n (%) | | | | |
| BCD | 92 (80.70%) | 29 (65.91%) | 46 (77.97%) | 1,078 (70.87%) |
| NTZ | 22 (19.30%) | 15 (34.09%) | 13 (22.03%) | 443 (29.13%) |
| Baseline Total Healthcare Utilization: number of codes and CUIs, mean (SD) | 1403.82 (1572.31) | 1584.34 (1280.30) | 1548.83 (1315.00) | 1359.05 (1512.99) |
| Baseline PDDS Risk: mean (SD) | 1.83 (0.65) | 1.83 (0.53) | 2.23 (0.81) | 2.07 (0.71) |
| Year 1 PDDS Risk: mean (SD) | 1.73 (0.77) | 1.64 (0.53) | 2.29 (1.05) | 2.13 (0.91) |

Cohort characteristics with mean (standard deviation) were presented for continuous variables and count (percentage) were presented for categorical variables. Baseline total healthcare utilization corresponds to the count of all codified EHR features and narrative CUI features observed during distinct clinical encounters occurring prior to target treatment initiation. Baseline PDDS risk was derived as the predicted mean PDDS as time of target treatment initiation, using independently fitted PDDS imputation models. Year 1 PDDS risk was calculated as the average of observed PDDS scores within one year of target treatment initiation for observations with available PDDS data and otherwise imputed by the same, independent PDDS imputation models using additional covariate information through one-year following target treatment initiation.

**Abbreviations**: BCD, B-cell depletion. CUI, Concept Unique Identifier. NTZ, Natalizumab. PDDS, Patient Determined Disease Steps.

### eTable 4b. Cohort Characteristics (by 3-Year Evaluation Window)

| Variable | No Sustained Change (n=130) | Sustained Improvement (n=92) | Sustained Worsening (n=118) | Unlabeled (n=1398) |
|---|---|---|---|---|
| Age at Target Treatment Initiation: years, mean (SD) | 41.30 (13.43) | 44.04 (11.85) | 47.16 (12.61) | 46.44 (13.01) |
| Self-Reported Gender: n (%) | | | | |
| Men | 37 (28.46%) | 16 (17.39%) | 38 (32.20%) | 397 (28.40%) |
| Women | 93 (71.54%) | 76 (82.61%) | 80 (67.80%) | 1,001 (71.60%) |
| Race and ethnicity: n (%) | | | | |
| White, Non-Hispanic | 110 (84.62%) | 82 (89.13%) | 105 (88.98%) | 1,210 (86.55%) |
| Other race and ethnic groups | 20 (15.38%) | 10 (10.87%) | 13 (11.02%) | 188 (13.45%) |
| Disease Duration: years, mean (SD) | 4.49 (4.91) | 4.05 (4.46) | 5.58 (5.51) | 5.38 (4.69) |
| Follow-up Duration: years, mean (SD) | 9.75 (4.88) | 9.85 (4.65) | 9.79 (4.82) | 9.20 (5.03) |
| Target Treatment Class: n (%) | | | | |
| BCD | 105 (80.77%) | 64 (69.57%) | 88 (74.58%) | 988 (70.67%) |
| NTZ | 25 (19.23%) | 28 (30.43%) | 30 (25.42%) | 410 (29.33%) |
| Baseline Total Healthcare Utilization: number of codes and CUIs, mean (SD) | 1264.22 (1182.96) | 1529.38 (1277.17) | 1528.79 (1581.96) | 1361.09 (1538.16) |
| Baseline PDDS Risk: mean (SD) | 1.94 (0.74) | 1.93 (0.62) | 2.19 (0.72) | 2.07 (0.71) |
| Year 1 PDDS Risk: mean (SD) | 1.88 (0.95) | 1.81 (0.70) | 2.21 (0.93) | 2.13 (0.91) |

Cohort characteristics with mean (standard deviation) were presented for continuous variables and count (percentage) were presented for categorical variables. Baseline total healthcare utilization corresponds to the count of all codified EHR features and narrative CUI features observed during distinct clinical encounters occurring prior to target treatment initiation. Baseline PDDS risk was derived as the predicted mean PDDS as time of target treatment initiation, using independently fitted PDDS imputation models. Year 1 PDDS risk was calculated as the average of observed PDDS scores within one year of target treatment initiation for observations with available PDDS data and otherwise imputed by the same, independent PDDS imputation models using additional covariate information through one-year following target treatment initiation.
**Abbreviations**: BCD, B-cell depletion. CUI, Concept Unique Identifier. NTZ, Natalizumab. PDDS, Patient Determined Disease Steps.

### eTable 4c. Cohort Characteristics (by Target Treatment Group)

| Variable | BCD (n=1245) | NTZ (n=493) |
|---|---|---|
| Age at Target Treatment Initiation: years, mean (SD) | 47.33 (13.39) | 42.56 (11.37) |
| Self-Reported Gender: n (%) | | |
| Men | 394 (31.65%) | 94 (19.07%) |
| Women | 851 (68.35%) | 399 (80.93%) |
| Race and ethnicity: n (%) | | |
| White, Non-Hispanic | 1,073 (86.18%) | 434 (88.03%) |
| Other race and ethnic groups | 172 (13.82%) | 59 (11.97%) |
| Disease Duration: years, mean (SD) | 5.62 (4.84) | 4.33 (4.43) |
| Follow-up Duration: years, mean (SD) | 9.82 (4.95) | 8.05 (4.86) |
| Baseline Total Healthcare Utilization: number of codes and CUIs, mean (SD) | 1561.60 (1606.35) | 900.72 (1076.00) |
| Baseline PDDS Risk: mean (SD) | 2.13 (0.72) | 1.88 (0.66) |
| Year 1 PDDS Risk: mean (SD) | 2.14 (0.87) | 2.00 (0.98) |
| Post-Treatment Disability Outcome, n (%) | | |
| No Sustained Change | 105 (8.43%) | 25 (5.07%) |
| Sustained Improvement | 64 (5.14%) | 28 (5.68%) |
| Sustained Worsening | 88 (7.07%) | 30 (6.09%) |
| No Labeled Outcome | 988 (79.36%) | 410 (83.16%) |

Cohort characteristics with mean (standard deviation) were presented for continuous variables and count (percentage) were presented for categorical variables. Baseline total healthcare utilization corresponds to the count of all codified EHR features and narrative CUI features observed during distinct clinical encounters occurring prior to target treatment initiation. Baseline PDDS risk was derived as the predicted mean PDDS as time of target treatment initiation, using independently fitted PDDS imputation models. Year 1 PDDS risk was calculated as the average of observed PDDS scores within one year of target treatment initiation for observations with available PDDS data and otherwise imputed by the same, independent PDDS imputation models using additional covariate information through one-year following target treatment initiation.

**Abbreviations**: BCD, B-cell depletion. CUI, Concept Unique Identifier. NTZ, Natalizumab. PDDS, Patient Determined Disease Steps.

### eTable 5a. Cohort Characteristics by Target Treatment Group: Comparing Observed and DiPS-Balanced Statistics (Sustained Improvement Outcome)

| Variable | BCD (n=1245) | NTZ (n=493) | P-Value | BCD (n=1245) | NTZ (n=493) | P-Value |
|---|---|---|---|---|---|---|
| | Observed, Unbalanced | | | Balanced by DiPS (Sustained Improvement-Informed) | | |
| Age at Target Treatment Initiation: years, mean (SD) | 47.33 (13.39) | 42.56 (11.37) | <0.001 | 46.03 (13.51) | 44.59 (11.84) | 0.092 |
| Self-Reported Gender: n (%) | | | | | | |
| Men | 394 (31.65%) | 94 (19.07%) | <0.001 | 470 (27.85%) | 343 (23.22%) | 0.028 |
| Women | 851 (68.35%) | 399 (80.93%) | | 1,217 (72.15%) | 1,133 (76.78%) | |
| Race and ethnicity: n (%) | | | | | | |
| White, Non-Hispanic | 1,073 (86.18%) | 434 (88.03%) | 0.345 | 1,461 (86.65%) | 1,315 (89.12%) | 0.118 |
| Other race and ethnic groups | 172 (13.82%) | 59 (11.97%) | | 225 (13.35%) | 161 (10.88%) | |
| Disease Duration: years, mean (SD) | 5.62 (4.84) | 4.33 (4.43) | <0.001 | 5.19 (4.74) | 5.08 (4.68) | 0.745 |
| Follow-up Duration: years, mean (SD) | 9.82 (4.95) | 8.05 (4.86) | <0.001 | 9.34 (5.09) | 9.23 (5.00) | 0.601 |
| Baseline Total Healthcare Utilization: number of codes and CUIs, mean (SD) | 1561.60 (1606.35) | 900.72 (1076.00) | <0.001 | 1392.18 (1479.45) | 1327.13 (1513.32) | 0.054 |
| Baseline PDDS Risk: mean (SD) | 2.13 (0.72) | 1.88 (0.66) | <0.001 | 2.06 (0.71) | 2.00 (0.68) | 0.316 |

Cohort characteristics with mean (standard deviation) were presented for continuous variables and count (percentage) were presented for categorical variables. P-values for results after balancing were reported from weighted chi-square tests for categorical variables (race-ethnicity and gender) and weighted Mann-Whitney U-tests for the remaining, continuous variables. P-values for observed (i.e., unbalanced) characteristics were calculated using unweighted versions of the same tests. The subset of variables from Table 1 defined only by pre-target treatment initiation were included. Only the difference in self-reported gender remains statistically significant, but the observed difference between treatment groups was modest (4.6 percentage points) and showed largely improved covariate balance to the observed proportions. Baseline total healthcare utilization corresponded to the count of all codified EHR features and narrative CUI features observed during distinct clinical encounters occurring prior to target treatment initiation. Baseline PDDS risk was derived as the predicted mean PDDS as time of target treatment initiation, using independently fitted PDDS imputation models. Year 1 PDDS risk was calculated as the average of observed PDDS scores within one year of target treatment initiation for observations with available PDDS data and otherwise imputed by the same, independent PDDS imputation models using additional covariate information through one-year following target treatment initiation.
**Abbreviations**: BCD, B-cell depletion. CUI, Concept Unique Identifier. NTZ, Natalizumab. PDDS, Patient Determined Disease Steps.

**eTable 5b. Cohort Characteristics by Target Treatment Group: Comparing Observed and DiPS-Balanced Statistics (Sustained Worsening Outcome)**

| Variable | BCD (n=1245) | NTZ (n=493) | P-Value | BCD (n=1245) | NTZ (n=493) | P-Value |
|---|---|---|---|---|---|---|
| | Observed, Unbalanced | | | Balanced by DiPS (Sustained Worsening-Informed) | | |
| Age at Target Treatment Initiation: years, mean (SD) | 47.33 (13.39) | 42.56 (11.37) | <0.001 | 45.93 (13.54) | 45.08 (12.49) | 0.325 |
| Self-Reported Gender: n (%) | | | <0.001 | | | |
| Men | 394 (31.65%) | 94 (19.07%) | | 474 (28.05%) | 350 (22.59%) | 0.009 |
| Women | 851 (68.35%) | 399 (80.93%) | | 1,217 (71.95%) | 1,198 (77.41%) | |
| Race and ethnicity: n (%) | | | 0.345 | | | |
| White, Non-Hispanic | 1,073 (86.18%) | 434 (88.03%) | | 1,465 (86.64%) | 1,373 (88.67%) | 0.198 |
| Other race and ethnic groups | 172 (13.82%) | 59 (11.97%) | | 226 (13.36%) | 175 (11.33%) | |
| Disease Duration: years, mean (SD) | 5.62 (4.84) | 4.33 (4.43) | <0.001 | 5.18 (4.74) | 5.38 (5.02) | 0.797 |
| Follow-up Duration: years, mean (SD) | 9.82 (4.95) | 8.05 (4.86) | <0.001 | 9.29 (5.11) | 9.34 (5.05) | 0.970 |
| Baseline Total Healthcare Utilization: number of codes, mean (SD) | 1561.60 (1606.35) | 900.72 (1076.00) | <0.001 | 1389.27 (1486.29) | 1409.20 (1544.53) | 0.345 |
| Baseline PDDS Risk: mean (SD) | 2.13 (0.72) | 1.88 (0.66) | <0.001 | 2.06 (0.71) | 2.05 (0.71) | 0.944 |

Cohort characteristics with mean (standard deviation) were presented for continuous variables and count (percentage) were presented for categorical variables. P-values were reported from weighted chi-square tests for categorical variables (race-ethnicity and gender) and weighted Mann-Whitney U-tests for the remaining, continuous variables. P-values for observed (i.e. unbalanced) characteristics were calculated using unweighted versions of the same tests. The subset of variables from Table 1 defined only by pre-target treatment initiation were included. As in eTable 5A, the difference in self-reported gender remained statistically significant, but the observed difference between treatment groups was similarly small (5.5 percentage points) and improved over the unbalanced proportions.
Baseline total healthcare utilization corresponded to the count of all codified EHR features and narrative CUI features observed during distinct clinical encounters occurring prior to target treatment initiation. Baseline PDDS risk was derived as the predicted mean PDDS as time of target treatment initiation, using independently fitted PDDS imputation models. Year 1 PDDS risk was calculated as the average of observed PDDS scores within one year of target treatment initiation for observations with available PDDS data and otherwise imputed by the same, independent PDDS imputation models using additional covariate information through one-year following target treatment initiation.
**Abbreviations**: BCD, B-cell depletion. CUI, Concept Unique Identifier. NTZ, Natalizumab. PDDS, Patient Determined Disease Steps.

### eTable 6a. Sensitivity Analysis Results for Sustained Worsening Models

| Endpoint | Model | ATE | Std. Err | 95% CI |
|---|---|---|---|---|
| **2-year** | Main Analysis | -0.013 | 0.037 | (-0.069, 0.074) |
| | All Analytic Features | 0.039 | 0.048 | (-0.095, 0.093) |
| | 95% Trim | -0.02 | 0.036 | (-0.067, 0.076) |
| | 3-month Sustainment Window | -0.018 | 0.043 | (-0.127, 0.047) |
| | 6-month Lookback | -0.016 | 0.039 | (-0.072, 0.087) |
| | 12-month Lookback | -0.04 | 0.041 | (-0.088, 0.075) |
| | Including DMT switching | 0.005 | 0.033 | (-0.062, 0.069) |
| **3-year** | Main Analysis | -0.020 | 0.058 | (-0.149, 0.076) |
| | All Analytic Features | 0.001 | 0.052 | (-0.125, 0.085) |
| | 95% Trim | -0.025 | 0.049 | (-0.124, 0.083) |
| | 3-month Sustainment Window | -0.033 | 0.060 | (-0.205, 0.031) |
| | 6-month Lookback | -0.016 | 0.078 | (-0.252, 0.048) |
| | 12-month Lookback | -0.022 | 0.073 | (-0.231, 0.051) |
| | Including DMT Switching | 0.005 | 0.047 | (-0.12, 0.074) |

**Abbreviations**: ATE, Average Treatment Effect. CI, Confidence Interval. DMT, Disease-Modifying Therapy Std. Err, Standard Error.

### eTable 6b. Sensitivity Analysis Results for Sustained Improvement Models

| Endpoint | Sensitivity Model | ATE | Std. Err | 95% CI |
|---|---|---|---|---|
| **2-year** | Main | -0.187 | 0.072 | (-0.264, 0.008) |
| | All Analytic Features | -0.151 | 0.076 | (-0.245, 0.047) |
| | 95% Trim | -0.147 | 0.071 | (-0.253, 0.017) |
| | 3-month Sustainment Window | -0.014 | 0.040 | (-0.095, 0.065) |
| | 6-month Lookback | -0.215 | 0.086 | (-0.289, 0.01) |
| | 12-month Lookback | -0.152 | 0.07 | (-0.251, 0.005) |
| | Including DMT Switching | -0.138 | 0.057 | (-0.203, 0.015) |
| **3-year** | Main | -0.073 | 0.052 | (-0.187, 0.009) |
| | All Analytic Features | -0.085 | 0.055 | (-0.177, 0.045) |
| | 95% Trim | -0.107 | 0.048 | (-0.178, 0.014) |
| | 3-month Sustainment Window | -0.008 | 0.044 | (-0.091, 0.083 |
| | 6-month Lookback | -0.106 | 0.065 | (-0.211, 0.03) |
| | 12-month Lookback | -0.111 | 0.056 | (-0.212, 0.01) |
| | Including DMT Switching | -0.075 | 0.046 | (-0.167, 0.013) |

**Abbreviations**: ATE, Average Treatment Effect. CI, Confidence Interval. DMT, Disease-Modifying Therapy Std. Err, Standard Error.

### eTable 7. Summary of Treatment Effect Estimates

| Evaluation Window | Outcome | ATE | Std. Err | 95% CI | P-Value |
|---|---|---|---|---|---|
| 3-years | Sustained Worsening | -0.020 | 0.058 | (-0.149, 0.076) | 0.755 |
| | Sustained Improvement | -0.073 | 0.052 | (-0.187, 0.009) | 0.114 |
| 2-years | Sustained Worsening | -0.013 | 0.037 | (-0.069, 0.074) | 0.819 |
| | Sustained Improvement | -0.187 | 0.072 | (-0.264, 0.008) | 0.135 |

**Abbreviations**: ATE, Average Treatment Effect. CI, Confidence Interval. Std. Err, Standard Error.

### eTable 8a. Power Analysis of 3-year Evaluation Window Analyses

| Outcome | Model | ATE | 95% CI | Std. Err | MDE |
|---|---|---|---|---|---|
| Sustained Improvement | Labelled Data Only | -0.184 | (-0.229, 0.009) | 0.060 | 0.132 |
| | Semi-Supervised (3-month PDDS separation) | -0.008 | (-0.091, 0.083) | 0.044 | 0.096 |
| | Semi-Supervised (6-month PDDS separation) | -0.073 | (-0.187, 0.009) | 0.052 | 0.114 |
| Sustained Worsening | Labelled Data Only | 0.012 | (-0.129, 0.079) | 0.055 | 0.122 |
| | Semi-Supervised (3-month PDDS separation) | -0.033 | (-0.205, 0.031) | 0.060 | 0.133 |
| | Semi-Supervised (6-month PDDS separation) | -0.020 | (-0.149, 0.076) | 0.058 | 0.127 |

**Abbreviations**: ATE, Average Treatment Effect. CI, Confidence Interval. Std. Err, Standard Error. MDE, Minimum Detectable Effect. PDDS, Patient Determined Disease Steps.

### eTable 8b. Power Analysis of 2-year Evaluation Window Analyses

| Outcome | Model | ATE | 95% CI | Std. Err | MDE |
|---|---|---|---|---|---|
| Sustained Improvement | Labelled Data Only | -0.132 | (-0.288, -0.010) | 0.070 | 0.157 |
| | Semi-Supervised (3-month PDDS separation) | -0.014 | (-0.095, 0.065) | 0.040 | 0.087 |
| | Semi-Supervised (6-month PDDS separation) | -0.187 | (-0.264, 0.008) | 0.072 | 0.165 |
| Sustained Worsening | Labelled Data Only | 0.027 | (-0.099, 0.136) | 0.061 | 0.134 |
| | Semi-Supervised (3-month PDDS separation) | -0.018 | (-0.127, 0.047) | 0.043 | 0.089 |
| | Semi-Supervised (6-month PDDS separation) | -0.013 | (-0.069, 0.074) | 0.037 | 0.079 |

**Abbreviations**: ATE, Average Treatment Effect. CI, Confidence Interval. Std. Err, Standard Error. MDE, Minimum Detectable Effect. PDDS, Patient Determined Disease Steps.
